# Mapping intronic mutational hotspots by *in silico* mutagenesis enables single antisense oligonucleotide correction of multiple variants

**DOI:** 10.64898/2026.09.10.26362533

**Authors:** Hassan Saei, Béatrice Ardin, Nicolas Kaiser, Mouad Ouahmane, Olivier Gribouval, Vincent Moriniere, Manon Mautret-Godefroy, Florian J. Wopperer, Korbinian M. Riedhammer, Daniel P. Gale, Omid Sadeghi-Alavijeh, Claire Goursaud, Olivier Grunewald, Chloe Prosper, Louis Lebreton, Marion Rabant, Carsten Bergmann, Raphael Kormann, Stephane Decramer, Bertrand Knebelmann, Corinne Antignac, Michael S. Wiesener, Guillaume Dorval

## Abstract

Deep intronic variants remain an understudied class of pathogenic variation, primarily due to their absence from standard exome and gene panel datasets and the complexity of non-coding genome interpretation. We hypothesized that pathogenic deep intronic variants are not randomly distributed but instead cluster within intronic “hotspots” inherently prone to pseudoexon activation, and that mapping such regions could improve molecular diagnosis. Importantly, such intronic hotspots nominate targets for antisense oligonucleotide (ASO) therapy. Using X-linked Alport syndrome as a proof-of-concept model and based on our previous work, we screened unsolved patients across multiple European diagnostic centers for variants within a defined region of *COL4A5* intron 6. We identified eight independent variants in more than 35 affected individuals from ten unrelated families, which led to two pseudoexon inclusion events, both using the same strong cryptic splice donor site. In all, RNA sequencing and/or minigene assays confirmed aberrant splicing, even when prediction tools were discordant or fell below clinical thresholds. A single ASO targeting the shared donor site restored normal *COL4A5* mRNA and α5(IV) collagen protein expression in patient-derived cells regardless of the causative variant. Extending this analysis gene-wide using the AlphaGenome sequence-to-function model, we confirmed intron 6 as one of the most critical *COL4A5* splicing hotspot and identified additional potential hotspots harboring novel predicted spliceogenic variants. This gene-agnostic framework establishes a systematic strategy for identifying intronic mutational hotspots and matching patients to scalable, mutation-agnostic ASO-based precision therapies.

**Significance Statement:** Deep intronic variants are an underrecognized cause of genetic disease, undetectable to standard exome and gene-panel testing and difficult to interpret. Using X-linked Alport syndrome as an exemplar, we identified a recurrent deep intronic mutational hotspot in which independent pathogenic variants from unrelated families converge on the same cryptic splice donor site to activate pseudoexon inclusion. A single antisense oligonucleotide therapy corrected this shared splicing defect and restored normal collagen IV protein production in patient cells, regardless of the underlying causative variant. Extending this analysis gene-wide using an AlphaGenome sequence-to-function splicing model, we uncovered additional deep intronic hotspots across the gene, establishing a gene-agnostic strategy for systematically identifying hidden pathogenic variants and matching patients to mutation-agnostic, RNA-based precision therapies in Mendelian disorders.

## Introduction

Over the past two decades, massively parallel, or next generation, sequencing has transformed the molecular diagnosis of Mendelian disorders, enabling systematic identification of pathogenic variants in coding regions and canonical splice sites. Yet a substantial fraction of patients with a clear clinical diagnosis remain genetically unsolved after standard genomic evaluation, suggesting that disease-causing variants reside beyond the reach of conventional approaches. The widespread adoption of genome sequencing and transcriptome profiling by RNA sequencing now grants unprecedented access to the non-coding genome, revealing deep intronic variants as an underrecognized, and largely undercharacterized class of pathogenic variations (1–5). These variants are difficult to interpret, as conventional splicing prediction tools, while improving, still suffer from limited sensitivity in some non-coding regions. Identifying and characterizing deep intronic variants is therefore critical, both for improving diagnostic yield in unsolved cases and for guiding the development of targeted therapies.

Among emerging therapeutic strategies, antisense oligonucleotides (ASOs) have shown remarkable potential to correct pathogenic splicing defects at RNA level (6–9), with a growing body of clinical evidence supporting their use in genetically defined patient subgroups. A key prerequisite for such precision approaches, however, is the identification of recurrent variants or shared splicing mechanisms, ideally organized around mutational hotspots, that enable treatment strategies applicable to groups of patients rather than isolated families (10). Systematically mapping potential intronic mutational hotspots, defined as discrete genomic regions disproportionately prone to pathogenic splicing alterations, thus represents a dual priority: strengthening molecular diagnosis and delineating actionable therapeutic targets.

In this study, we used X-linked Alport syndrome (XLAS) as a proof-of-concept model. This condition is caused by pathogenic variants in *COL4A5*, encoding the α5 chain of the trimeric type IV collagen (collagen α5(IV)), an essential component of the mature glomerular basement membrane. Alport syndrome is the second most frequent hereditary cause of kidney disease. In severely affected hemizygouse males, XLAS is characterized by progressive hematuric kidney failure, with or without hearing impairment and eye abnormalities (11, 12). Increasing evidence indicates that deep-intronic variants represent an underrecognized cause of *COL4A5*-related disease by inducing aberrant pre-mRNA splicing all converging on a single cryptic splice donor site and all amenable to correction by a single splice-switching ASO. Building on our pioneering demonstration that pseudoexon-activating *COL4A5* variants can be corrected by ASOs (6), we report herein the identification of a mutational hotspot within intron 6 of *COL4A5*, harboring multiple distinct variants in over 35 affected individuals across European diagnostic centers, all converging on a single cryptic splice donor site and all amenable to correction by a single splice-switching ASO. We further leveraged AlphaGenome (19), a state-of-the-art sequence-to-function model, to validate this hotspot *in silico*, and systematically map actionable deep intronic loci across the entire *COL4A5* gene leading to the identification of five novel deep-intronic mutational hotspots. Together, these findings establish a scalable analytical framework for the interrogation of intronic hotspots, with immediate diagnostic and therapeutic implications for Alport syndrome and, more broadly, for any Mendelian disease gene.

## Results

### Cohort Screening Reveals a Deep Intronic Mutational Hotspot in *COL4A5*

In a previous study, we identified numerous deep intronic variations responsible for X-linked Alport syndrome in 19 patients (4). In particular, two families had variations within a restricted region in intron 6 (c.385-660A>G and c.385-714G>A).

A supervised search for variants in this region of interest within a cohort from Necker Hospital comprising 2,680 patients with suspected hereditary kidney diseases led to the identification of 10 new families with variants in the region of interest. Among them, three additional independent families carried the previously identified c.385-660A>G variant, and seven others carried six new variants (two independent families carried the c.385-756C>G variant) (**Fig. 1 – Table 1**). Among the six new variants identified in our cohort (**Table 1**), only c.385-756C>G had been previously reported (ASN 2021 abstract; PO1304). For all of these families, X-linked Alport syndrome was suspected based on the pedigree or even confirmed by a lack of α5-chain labeling on the skin biopsy (**Table 2**). In total, we identified more than 35 patients in 12 families studied at our center with a pathogenic or suspected to be pathogenic variant in this specific region of *COL4A5*.

**Fig. 1.**
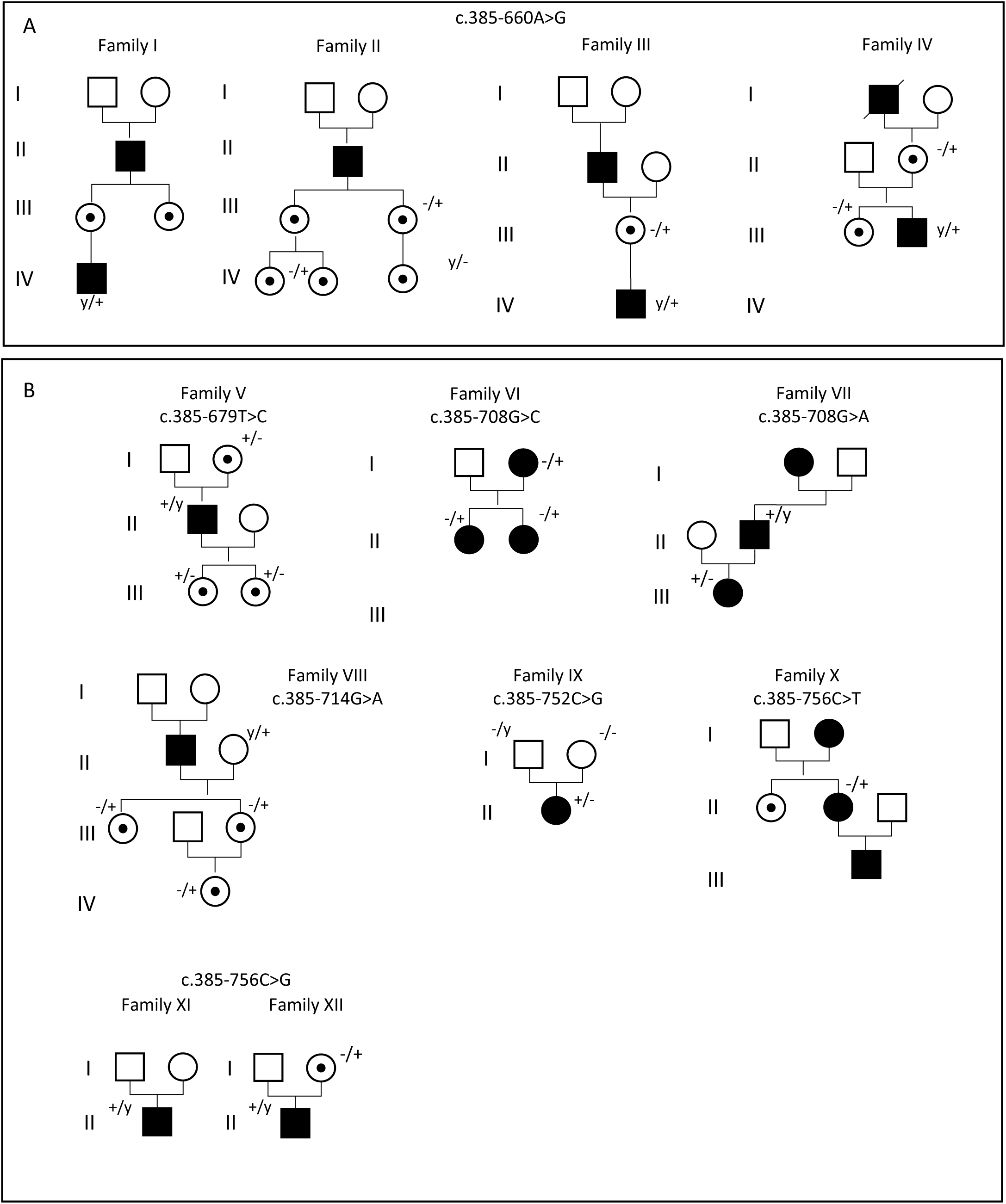
Pedigrees of patients with X-linked Alport syndrome carrying deep intronic variants in intron 6. (A) Four unrelated families from our local cohort were identified with the deep intronic variant c.385-660A>G. (B) Additional families identified locally carry distinct deep intronic variants within intron 6, all leading to the same pseudoexon activation event in this region. Females with hematuria are represented by a circle with a black dot in the middle, whereas a fully filled (plain black) circle corresponds to females that present not only hematuria but also further features of kidney disease. + indicates pathogenic variant; and –, no pathogenic variant. Clinical information is reported in Table 2. Familly III is published here (4). The square bracket in family XI indicate an adopted individual.

**Table 1.**
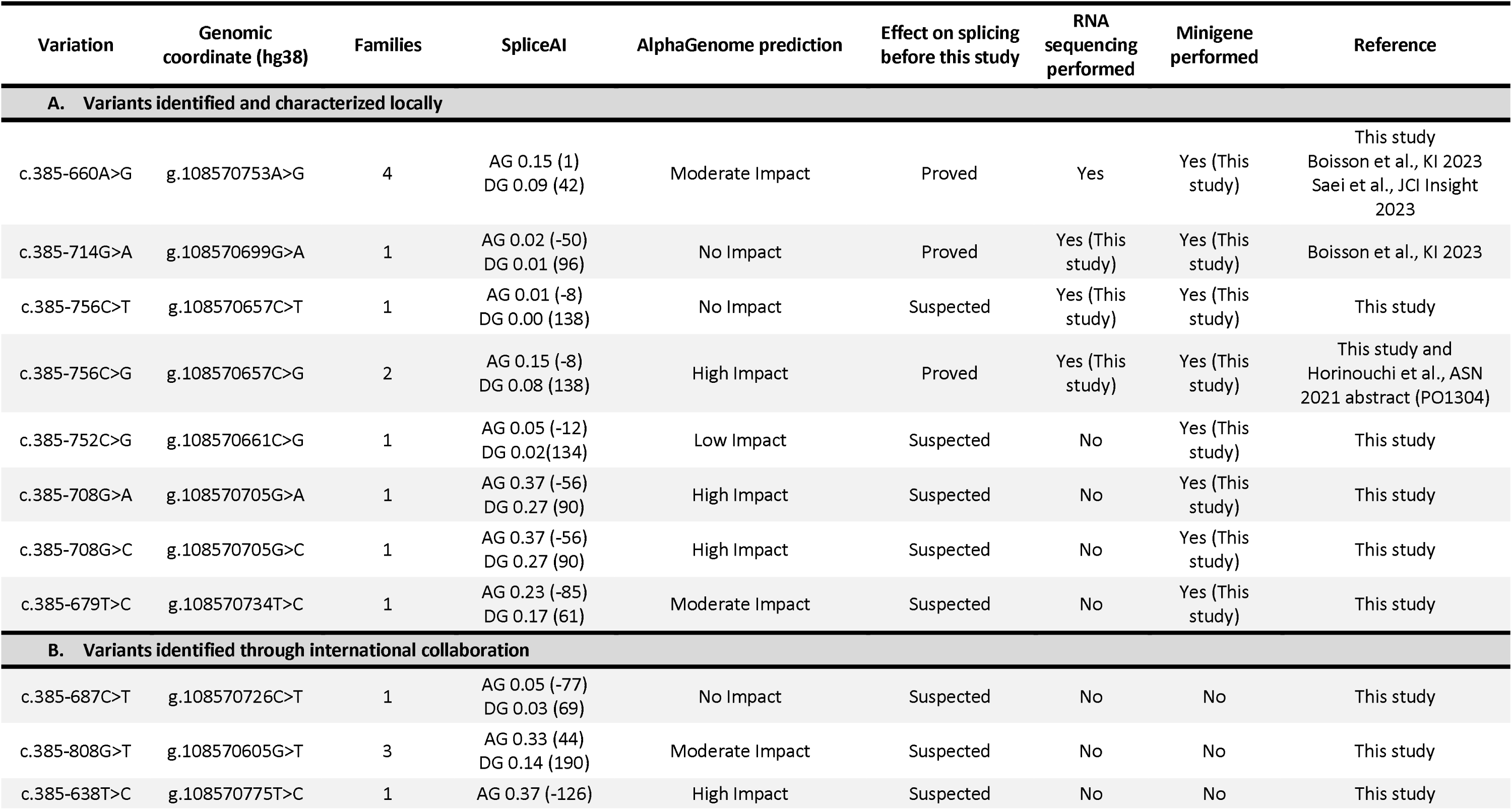
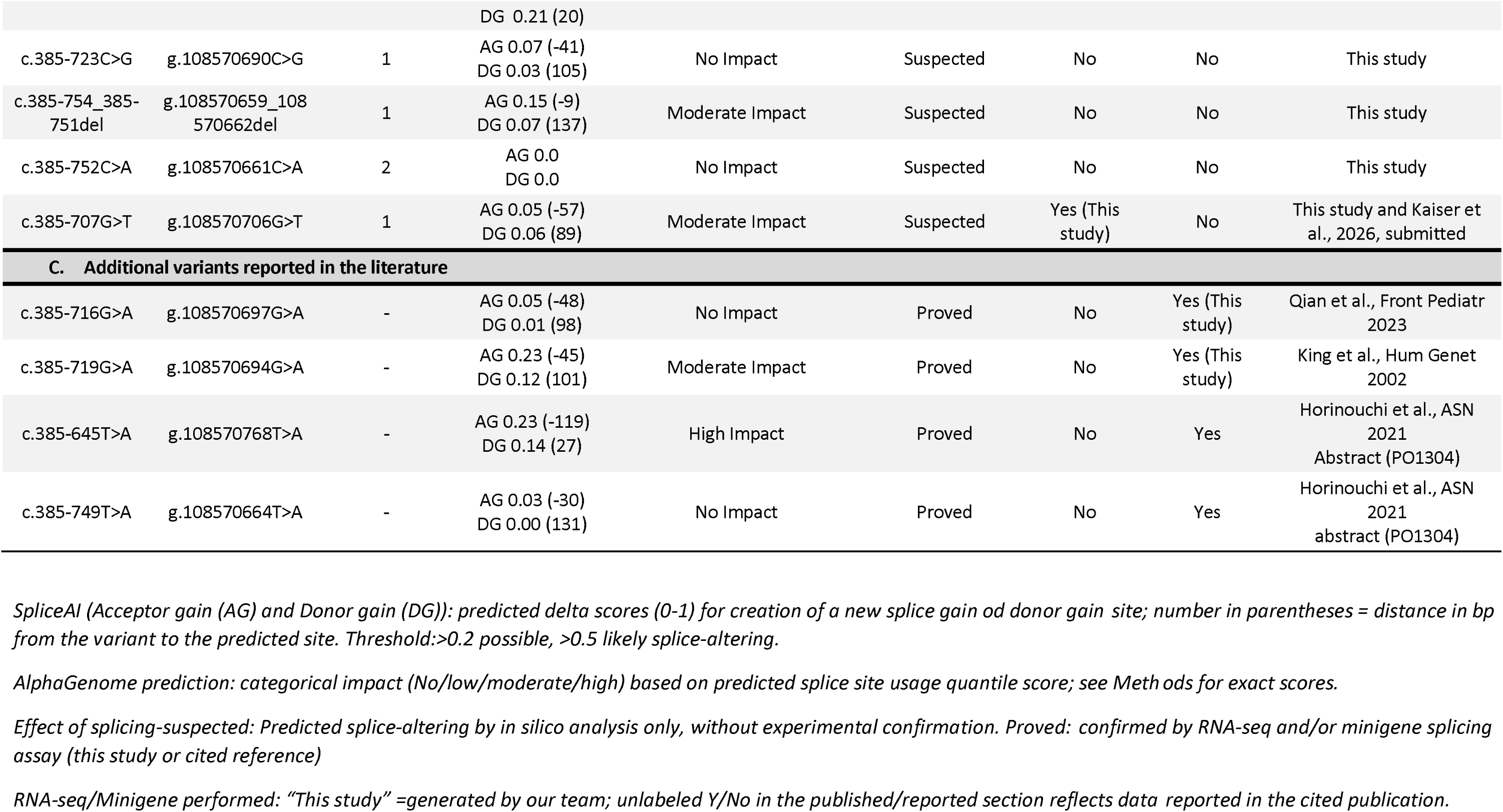
Variants identified and characterized in *COL4A5* intron 6 hotspot.

**Table 2.** Clinical characteristics of the confirmed intron 6 cohort.

| Family | Individual | Sex | Variation intron 6 |  | CKD stage | Age range | Hearing impairment | Ocular symptoms | skin biopsy |
| --- | --- | --- | --- | --- | --- | --- | --- | --- | --- |
| I | IV-2 | M | c.385-660A>G | Hemizygous | 1 | 0-5 | no | no |  |
| II | IV-2 | F | c.385-660A>G | Heterozygous | 1 | 36-40 | unknown | unknown |  |
| II | II-1 | M | c.385-660A>G | Hemizygous | 5 | 21-25 | yes | yes |  |
| II | IV-1 | F | c.385-660A>G | Heterozygous | 1 | 6-10 | no | no |  |
| II | IV-3 | F | not tested |  | 1 | 0-5 | no | no |  |
| III | II-2 | F | c.385-660A>G | Heterozygous | 2 | 61-65 | yes | no |  |
| III | III-3 | M | c.385-660A>G | Hemizygous | 4 | 26-30 | no | no | No staining |
| IV | III-2 | M | c.385-660A>G | Hemizygous | 5 | 26-30 | yes | no | No staining |
| IV | III-1 | F | c.385-660A>G | Heterozygous | 2 | 31-35 | yes | no | Segmental staining |
| IV | II-2 | F | c.385-660A>G | Heterozygous | 2 | 56-60 | yes | no |  |
| V | III-1 | F | c.385-679T>C | Heterozygous | 1 | 26-30 | no | no | Segmental staining |
| V | III-2 | F | c.385-679T>C | Heterozygous | 1 | 16-20 | no | no |  |
| V | II-1 | M | c.385-679T>C | Hemizygous | 5 | 36-40 | no | no |  |
| V | I-2 | F | c.385-679T>C | Heterozygous | 1 | 71-75 | no | no |  |
| VI | I-2 | F | c.385-708G>C | Heterozygous | 5 | 36-40 | no | no |  |
| VI | II-2 | F | c.385-708G>C | Heterozygous | 1 | 36-40 | yes | - | Segmental Staining |
| VI | II-3 | F | c.385-708G>C | Heterozygous | 1 | 31-35 | no | - |  |
| VII | III-1 | F | c.385-708G>A | Heterozygous | 1 | 16-20 | no | no |  |
| VII | II-2 | M | c.385-708G>A | Hemizygous | 5 | 26-30 | yes | - | No staining |
| VII | II-4 | F | not tested |  | 5 | 41-45 | no | no |  |
| VII | II-5 | F | c.385-708G>A | Heterozygous | 5 | 41-45 | no | no |  |
| VIII | II-3 | M | c.385-714G>A | Hemizygous | 3 | 71-75 | no | no |  |
| VIII | III-1 | F | c.385-714G>A | Heterozygous | 1 | 36-40 | no | no |  |
| VIII | IV | F | c.385-714G>A | Heterozygous | 1 | 41-45 | no | no |  |
| VIII | III-3 | F | c.385-714G>A | Heterozygous | 1 | 11-15 | no | no | Segmental staining |
| IX | II-2 | F | c.385-752C>G | Heterozygous | 1 | 11-15 | no | no |  |
| X | II-2 | F | c.385-756C>T | Heterozygous | 2 | 36-40 | no | no | Segmental staining |
| XI | II-1 | M | c.385-756C>G | Hemizygous | 1 | 11-15 | no | no | No staining |

### In Silico and Mechanistic Analyses Converge on a Shared Cryptic Donor Site Driving Two Pseudoexon-Inclusion Events

All variations were absent from population database (gnomAD). For each variant, we ran SpliceAI predicting tool (clinical threshold of 0.2) and recorded the maximum quantile score from each of AlphaGenome’s two splicing scorers (splice-junctions and splice-site-usage). This analysis revealed SpliceAI scores above clinical threshold in 3/8 variants whereas AlphaGenome reported variable splice site usage predictions (4/8 High Impact, 1/8 Moderate Impact, 1/8 Low Impact and 2/8 No Impact) (**Table 1** and **Fig. 2A-C**). AlphaGenome splicing scores for these variants are provided in **Table S1** and the predicted splice junctions (Sashimi plots) are shown in **Fig. S1**. All these variations were suspected to use the same donor splice site for the creation of two pseudoexons, one containing 42bp specific to the c.385-660A>G variation and the other of 147bp used by all other variations represented in **Fig. 2D**. The c.385-660A>G variation has been shown (4) to be responsible for the creation of a novel splicing acceptor site that does not exist in the reference sequence, whereas all other variants disrupt splice regulatory motifs (Intronic Splice Silencer ISS inactivation and/or Intronic Splice Enhancer creation, refer to **Table S2**), which activates the cryptic splice acceptor site at the position ChrX:108570648-108570649. Both pseudoexons contain an in-frame stop codon, which therefore leads to premature termination of translation, lacking the functionally critical C-terminal NC1 domain (which is necessary for trimer formation).

**Fig. 2.**
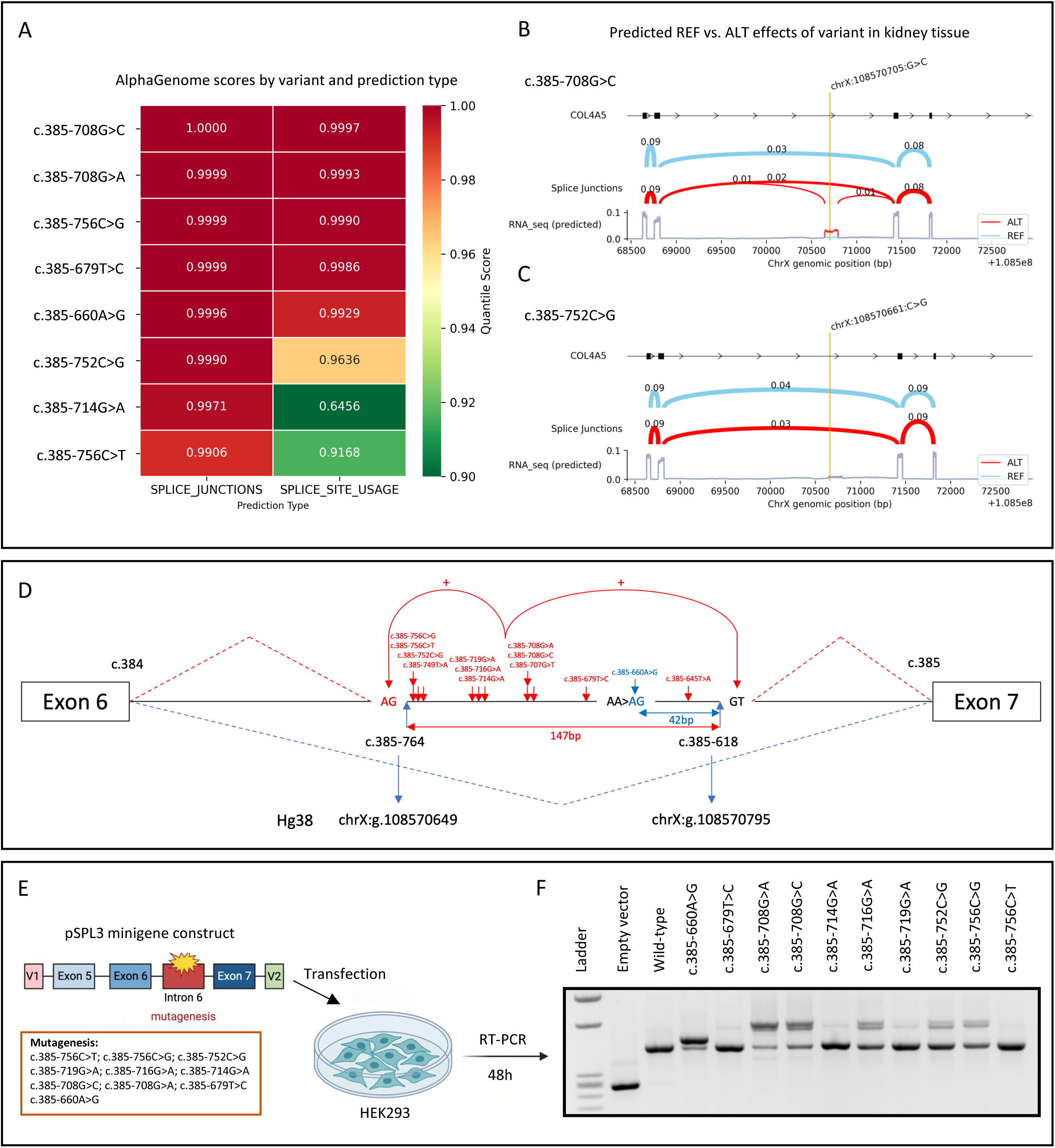
Characterization of deep intronic variants in intron 6 of the COL4A5 gene. (A) AlphaGenome analysis of splice junction and splice site usage for 8 variants identified in the local cohort within intron 6, where lower quantile scores indicate that the model failed to predict the variant impact on pseudoexon activation. (B) An example of variant with high predicted splice junction usage and (C) another for which AlphaGenome failed to predict pseudoexon activation. (D) Schematic representation of the intron 6 hotspot region showing all identified deep intronic variants, the resulting pseudoexon retention events, and the genomic coordinates defining the region of interest. (E, F) Functional assessment of intron 6 variants using a minigene splicing assay. Constructs containing exons 5, 6, intron 6 (541bp) and exon 7 of COL4A5 were cloned into the pSPL3 plasmid, transfected into HEK293 cells, and analyzed by RT-PCR after 48 hours.

### Functional Assays Validate Splicing Defects Even When In Silico Scores Are Weak

To demonstrate the effect of these variations, we conducted a minigene assay for all these variations and for two other previously published variations (17, 18) (**Fig. 2E and 2F**). The minigene assay showed clear evidence of abberent splicing for six out of 10 variants (strong pathogenic transcript). For the other four (c.385-679T>C, c.385-714G>A, c.385-719G>A, and c.385-756C>T), the pathogenic band was weak (but completely absent in the control). In all cases, including the four with a weak band, Sanger sequencing confirmed the sequences predicted *in silico* (42 and 147bp, respectively) (**Fig. S2**). Additionally, for three of the four variants with a weak band, we confirmed the pathogenic effect via RNA-seq analysis of cultured patients fibroblasts (**Fig. S3**). For the remaining variant (c.385-679T>C), RNA-seq analysis is currently ongoing. These results demonstrate the pathogenic effect of these variants and the presence of a mutational hotspot. Furthermore, while AlphaGenome outperformed SpliceAI in sensitivity in this region (6/8 vs. 3/8 variant predicted to be spliceogenic) (**Fig. 2B and C**), both tools showed discrepancies with functional results **(Fig. 2F**), suggesting that *in silico* scores alone are currently insufficient to support variant interpretation, and that experimental validation is required.

Upon identifying an intronic mutation hotspot, we sought to determine whether other research teams had also identified variations in this region. This led to the identification of 7 additional variations absent from the public gnomAD database in 10 new unrelated families with hematuric nephropathy consistent with XLAS (**Table 1 – Panel B**). Once again, *in silico* predictions in this cohort favored AlphaGenome over SpliceAI: only 2 of 7 variants exceeded the SpliceAI clinical threshold (>0.2), whereas AlphaGenome flagged 4 of 7 as spliceogenic (1 High Impact, 3 Moderate Impact. Notably, the two variants identified by SpliceAI (c.385-638T>C and c.385-808G>T) were also captured by AlphaGenome, showing concordance between the tools where SpliceAI did detect splicing impact. For one variant (c.385-707G>T) with non-significant SpliceAI score but moderate impact AlphaGenome prediction, the effect on splicing was confirmed by RT-PCR and RNA-seq on patient urine-derived renal epithelial cells (URECs) with and without nonsense-mediated decay (NMD) inhibition and resulted in the same 147bp retention in intron six of *COL4A5* (**Fig. 3A-D**). These results further supporting AlphaGenome’s improved sensitivity over SpliceAI.

**Fig. 3.**
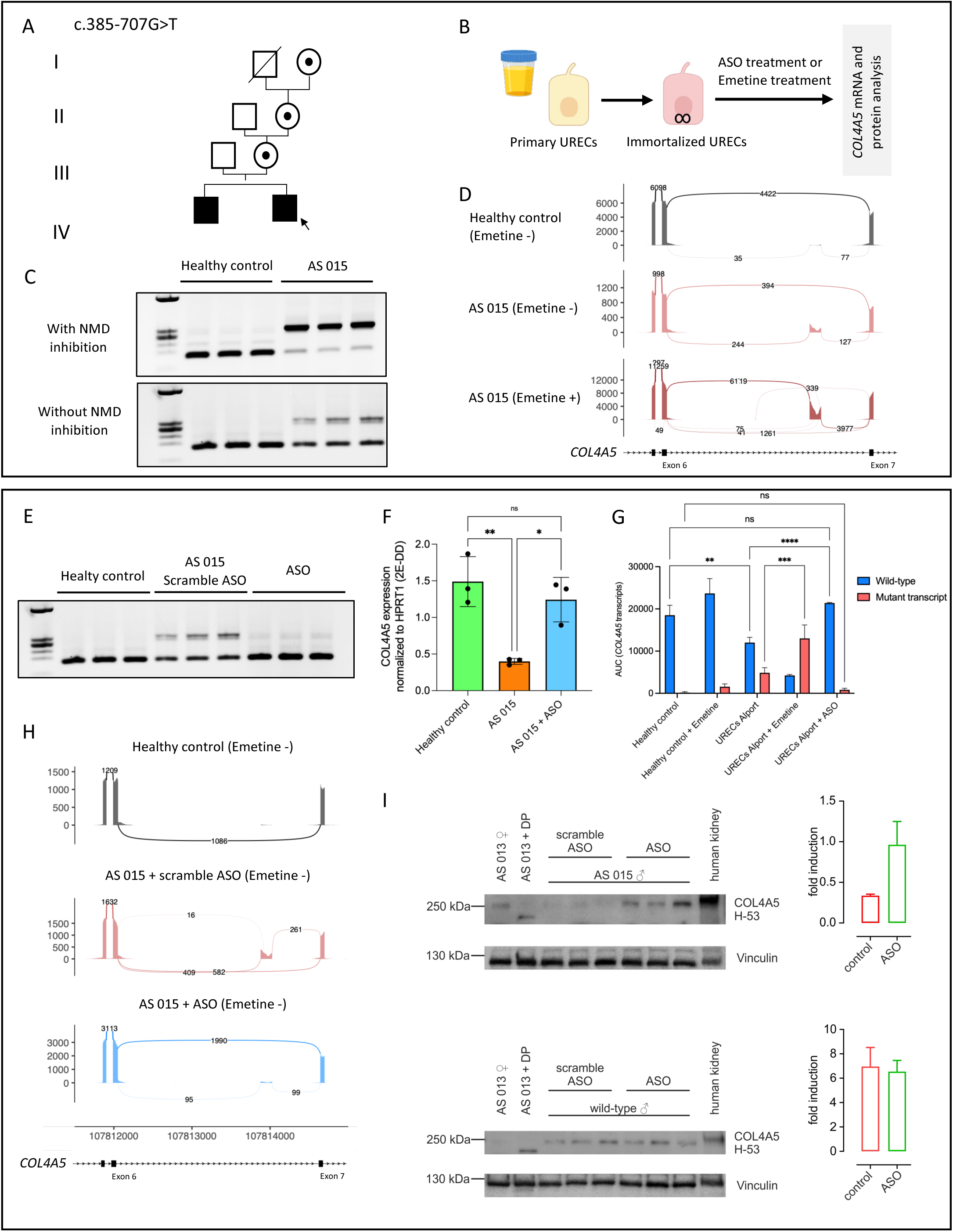
Characterization of COL4A5 mRNA and protein expression in patient-derived urinary epithelial cells carrying c.385-707G>T variant following ASO treatment. (A) Pedigree of the family carrying the COL4A5 c.385-707G>T variant (B) Schematic overview of sample collection from healthy donors and the Alport patient. (C) Qualitative RT-PCR analysis showing COL4A5 transcript expression levels in healthy control and Alport patient (AS015) URECs with and without the NMD inhibitor (emetine) treatment. (D) Sashimi plots showing targeted RNA-seq results in control and patient cells. (E) RT-PCR results showing COL4A5 expression in AS015 cells following ASO treatment. (F) RT-qPCR quantification showing COL4A5 gene expression before and after ASO treatment. (G) Fragment analyzer results showing the area under the curve for wild-type and mutant COL4A5 transcripts before and after ASO treatment. (H) Sashimi plots showing splice junctions across the intron 6 hotspot region before and after ASO treatment. (I) Western blot analysis showing collagen □5(IV) protein expression in URECs from the patient (AS 015) and his carrier mother (AS 013) with and without dipyridyl (DP) treatment. Collagen □5(IV) signal intensity was normalized to the vinculin control and shown relative to the signal intensity of untreated cells of AS 013 (=1.0). Data are presented as mean ± SEM. Statistical comparison was performed using the Mann-Whitney U test (p = 0.1).

### Single ASO Corrects Two Distinct Pseudoexon-Inclusion Events at a Shared Splice Donor site

In a previous study, our group demonstrated that the use of ASO specific to the splice donor site, when applied in the presence of the c.385-660A>G variant, inhibited the formation of the 42bp pseudoexon and thereby restored physiological splicing of *COL4A5* (6). Since all variants identified in this region share the same splice donor site, we sought to demonstrate that the oligonucleotides could restore splicing regardless of the variant and its effect (creation of a 42bp or 147bp pseudoexon). We thus studied the effect of the ASO on the c.385-707G>T variant, which causes the formation of a 147 pseudoexon.

URECs were subsequently treated with scramble and the functional ASO to mask the cryptic splice donor site utilized in both aberrant splicing events (**Fig. 3E**). Splice-switching treatment significantly increased *COL4A5* expression (p= 0.018) to the level of the control and fragment analysis confirmed a pronounced increase in wild-type transcripts (p<.0001) (**Fig. 3F, 3G and Fig. S4**). RNA sequencing further validated increased usage of the normal exon 6–7 junction following ASO treatment, with the ratio of wild-type to total (wild-type plus mutant) junctions reaching 95.26% after treatment (**Fig. 3H**). Western blot analysis of collagen α5(IV) protein expression after ASO treatment revealed an increased collagen α5(IV) signal intensity, reaching a level similar to the heterozygous mother, though not significantly different from that of the cells treated with a scramble ASO (p= 0.1) (**Fig. 3I**). No such effects were observed following ASO treatment of cells from a healthy proband.

### Systematic Unbiased Mutagenesis Identifies Potential Hotspot Introns Across *COL4A5*

To confirm intron 6 mutational hotspot and identify new hotspots in *COL4A5* intronic regions, we defined spliceogenic potential of all single-nucleotide substitutions across the entire *COL4A5* introns through saturation *in silico* mutagenesis (ISM) using AlphaGenome (52 introns, total variants analyzed=753348). For each intron, every possible substitution was scored for predicted splice site and splice junction usage, and variants were classified as High, Moderate, Low Impact and No Impact as described in the methods. High and Moderate Impact (H/M) variants were detected in nearly all introns but their number varied substantially with intron length and their positions in the intron (**Fig. 4A**). The largest total H/M impact counts were observed in intron 1 (407 variants), 6 (251), 30 (158), 41 (299), 42 (518), 43 (293), and 49 (130). Because a large fraction of H/M impact variants mapped close to canonical splice sites, we excluded for the analysis the first and last 100 nucleotides adjacent to exon-intron boundaries (**Fig. 4B**). This trimming revealed a more selective landscape of deep intronic spliceogenic variants, with prominent peaks remaining in intron 1 (365 H/M variants), 6 (151), 30 (123), 42 (138), 43 (78), 44 (76), and 49 (100) while many introns showed very few or no internal (deep intronic) spliceogenic variant.

**Fig. 4.**
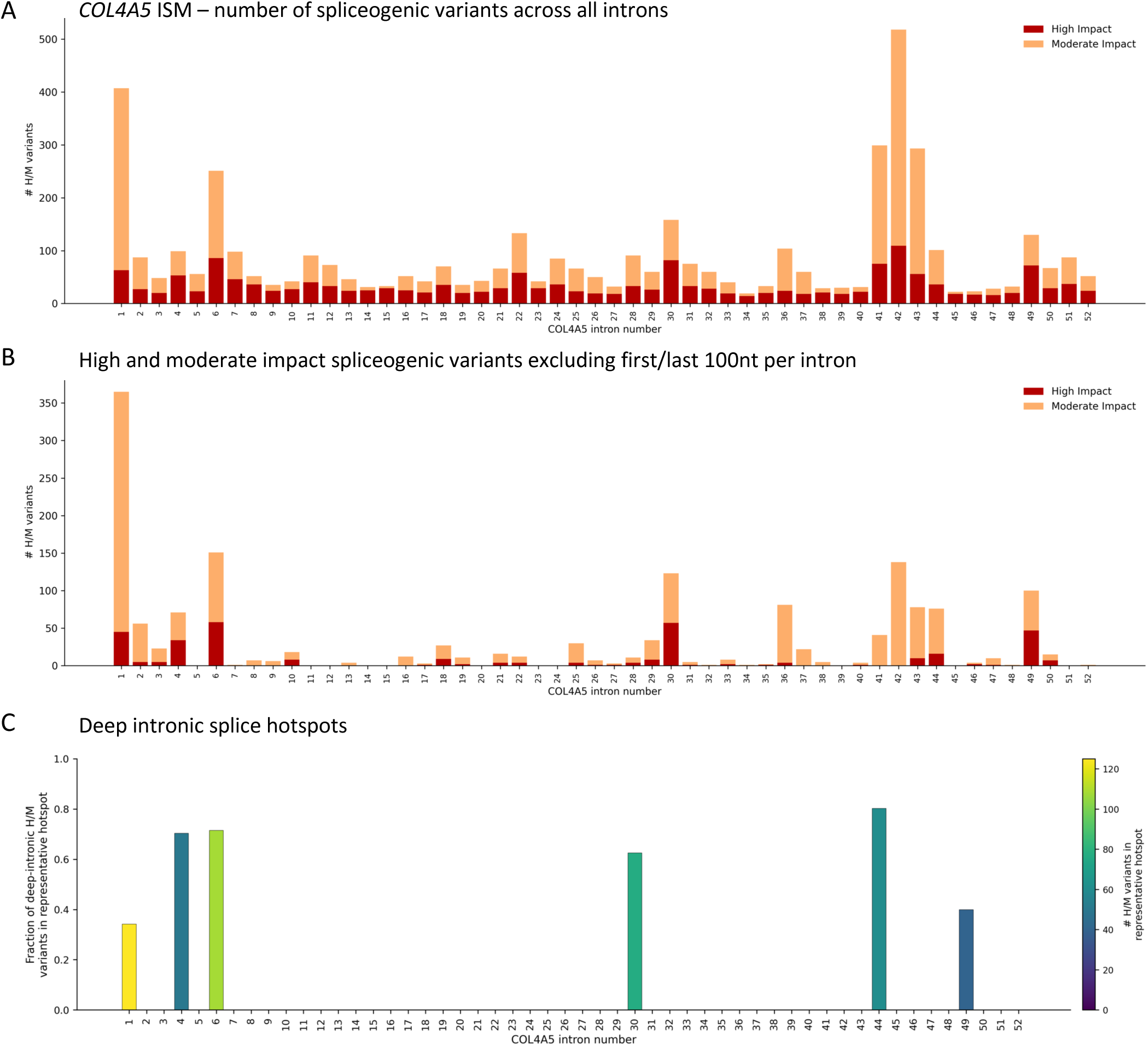
AlphaGenome ISM-predicted spliceogenic variant burden across COL4A5 introns identified deep intronic hotspots. (A) Total number of H/M variants per intron across the full intronic region. Dark red, High Impact; light orange, Moderate Impact. (B) Same as panel A, but restricted to the deep intronic region, defined as each intron excluding the first and last 100 nucleotides from the canonical splice sites flanking exons. This removes signal contamination by the spliceogenic substitutions from the canonical splice sites. (C) Hotspot across introns. Bar height shows the fraction of deep intronic H/M impact variants falling inside the significant hotspot interval defined by sliding-window significant enrichment analysis. Bar color indicates the number of H/M variants residing in those hotspots. Introns with high bar height and brighter color (e.g. introns 1, 4, 6, 30, 44, and 49) shows potential intron retention hotspots.

We next asked whether deep intronic H/M variants were randomly distributed or clustered into discrete intervals. Using 15-nucleotide sliding windows and Benjamini–Hochberg–adjusted enrichment testing (FDR < 0.05), we identified six introns with significant hotspot intervals: 1, 4, 6, 30, 44, and 49 (**Fig. 4C, Table S4**). Interestingly, intron 6 showed one of the strongest internal hotspots for H/M variants, with a high concentration of H/M variants 71.5% (108/151) clustered in a single 160bp region on chromosome X (chrX:108570641–108570801), where we identified all the pathogenic splice variants described above. Another strong hotspot interval was found in intron 44 (61/76 H/M - 80.2% - in chrX: 108680178-108680314). We further identified six patients in our database carrying deep intronic variants within this region, three of whom presented a phenotype highly suggestive of Alport syndrome (**Table S5**). Among these three, AlphaGenome classified one variant (c.3943-445T>C) as High Impact and a second (c.3943-409T>C) as Moderate Impact, while the remaining variants across the cohort were scored as No Impact, including one (c.3943-460A>T) with a proven effect in our previous study (4). These findings further support the utility of this unbiased hotspot-identification approach for prioritizing candidate variants in unresolved cases, pending functional validation.

For hotspot introns, ISM heatmaps illustrated the spatial localization of spliceogenic variants (hotspots in introns 6 and 44, and in introns 1, 4, 30 and 49 in **Fig. 5** and **Fig. S6 repectively**). In the intron 6 hotspot, saturation ISM across 480 substitutions (chrX:108,570,633–108,570,792; region size = 160 nt) identified 32 (6.7%) as High Impact, 67 (14%) as Moderate Impact, 86 (17.9%) as Low impact, and 295 (61.5%) as No impact (**Fig. 5A**). Among Low impact variants, only 37/86 with Splice Site Max Quantile above 0.9670 showed intron retention on AlphaGenome-predicted Sashimi junction plot. Intron 44 hotspot, (chrX:108,680,178–108,680,314; region size = 137 nt; 411 substitutions) contained 11 (2.7%) High Impact, 50 (12.2%) Moderate Impact, and 51 (12.4%) Low Impact. We also showed ISM result across a 123bp window (360 substitutions) in intron 47 (chrX:108,683,908–108,684,030, containing cryptic splice acceptor and donor sites and activated by variant c.4217-2008A>G causing a previously identified 106nt retention) to visually compare non-hotspot region to the significant hotspots (intron 6 and 44). The complete list of substitutions and their splicing scores for all hotspots are available on Zenodo (https://zenodo.org/records/19854065). The list of High Impact substitutions for hotspot introns is available in **Tables S6-11**.

**Fig. 5.**
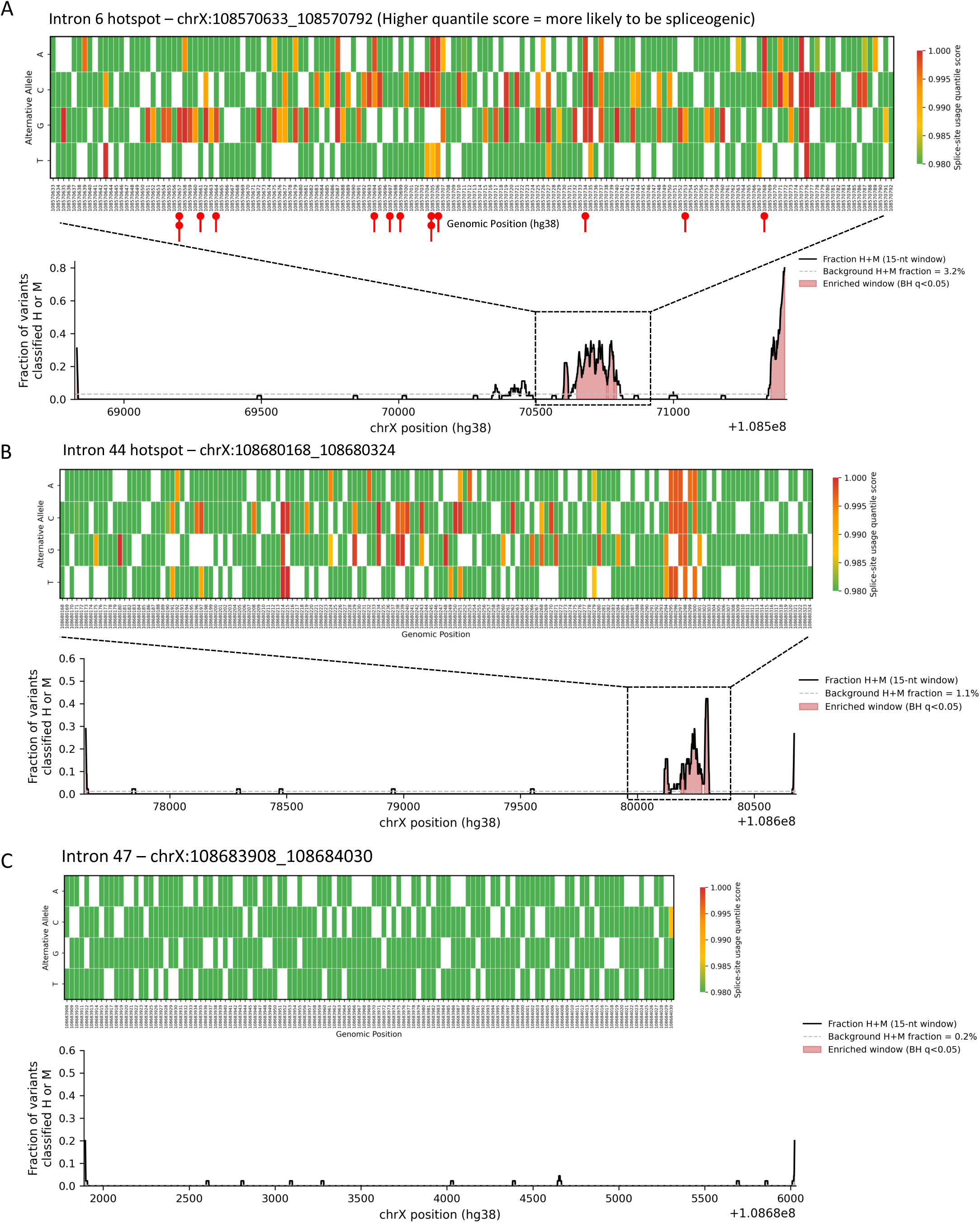
Representative spatial hotspot profiles in COL4A5 intron 6 and 44, compared with intron 47 as a negative control. (A) AlphaGenome ISM landscape across the 160nt deep-intronic window chrX:108,570,633–108,570,792 in intron 6. Each column corresponds to a single genomic position and each row to one of the three possible substitutions. Cells are colored by the AlphaGenome splice site usage quantile score (color scale 0.98 – 1.00; higher = more disruptive to splicing); white cells indicate the reference allele at that position. Variants predicted to most strongly disrupt splicing appear in red. Lollipops mark the 13 variants identified in Alport patients. (B) A narrow sharp enrichment peak is observed between (chrX:108680168-108680324) in intron 44 with tightly localized high-score cluster in the heatmap. After intron 6, this intron shows one of the highest hotspot concentrations in the analysis (∼89% of spliceogenic variants fall inside this region). (C) Representative 123nt deep-intronic window in intron 47, showing the low, non-enriched H/M scoring pattern (green) typical of most introns analyzed by ISM, in contrast to hotspot introns.

### Motif Enrichment Links Hotspot Spliceogenecity to Enhancer Gain, Silencer Loss and CG-Rich Motif Creation

To understand why substitutions in the six hotspot introns are predicted to be spliceogenic, we classified which trans-acting splicing factor binding sites were gained or lost in each mutant sequence compared to the reference, and which motifs were enriched among H/M impact mutant sequences using MSEA (**Fig. 6**). Across introns 1, 4, 6, 30, 44, and 49, H/M impact variants most often created or strengthened splicing enhancer motifs for SR proteins, rather than disrupting silencer elements (**Fig. 6A**). The most frequently gained motifs predicted to be for SF2/ASF protein, most prominent in intron 6 (34/99) and in intron 44 (26/62), SC35, strongest in intron 6 (27/99), and RESCUE-ESE hexamers, which were consistently gained across all six introns. Loss of the hnRNP A1 silencer motif was a secondary but reproducible signal in every intron, most frequently in intron 1 and 44. Consistent with this, rule-based mechanistic classification showed that splice enhancer gain was the dominant category in all six hotspot introns (**Fig. 6B**). A small subset of variants showed concurrent enhancer gain and hnRNP A1silencer motif loss, indicating a two-hit mechanism combining enhancer gain with silencer inactivation. Enhancer loss without a compensatory gain, and variants with no predicted motif change (”Unknown”), together accounted for the remainder, and were most prevalent in intron 1 (24.0% and 14.4%, respectively). Together, these results indicate that spliceogenic variants across the *COL4A5* intronic hotspots act predominantly through gain of SR-protein splicing enhancer motifs (**SI *Dataset 1***).

**Fig. 6.**
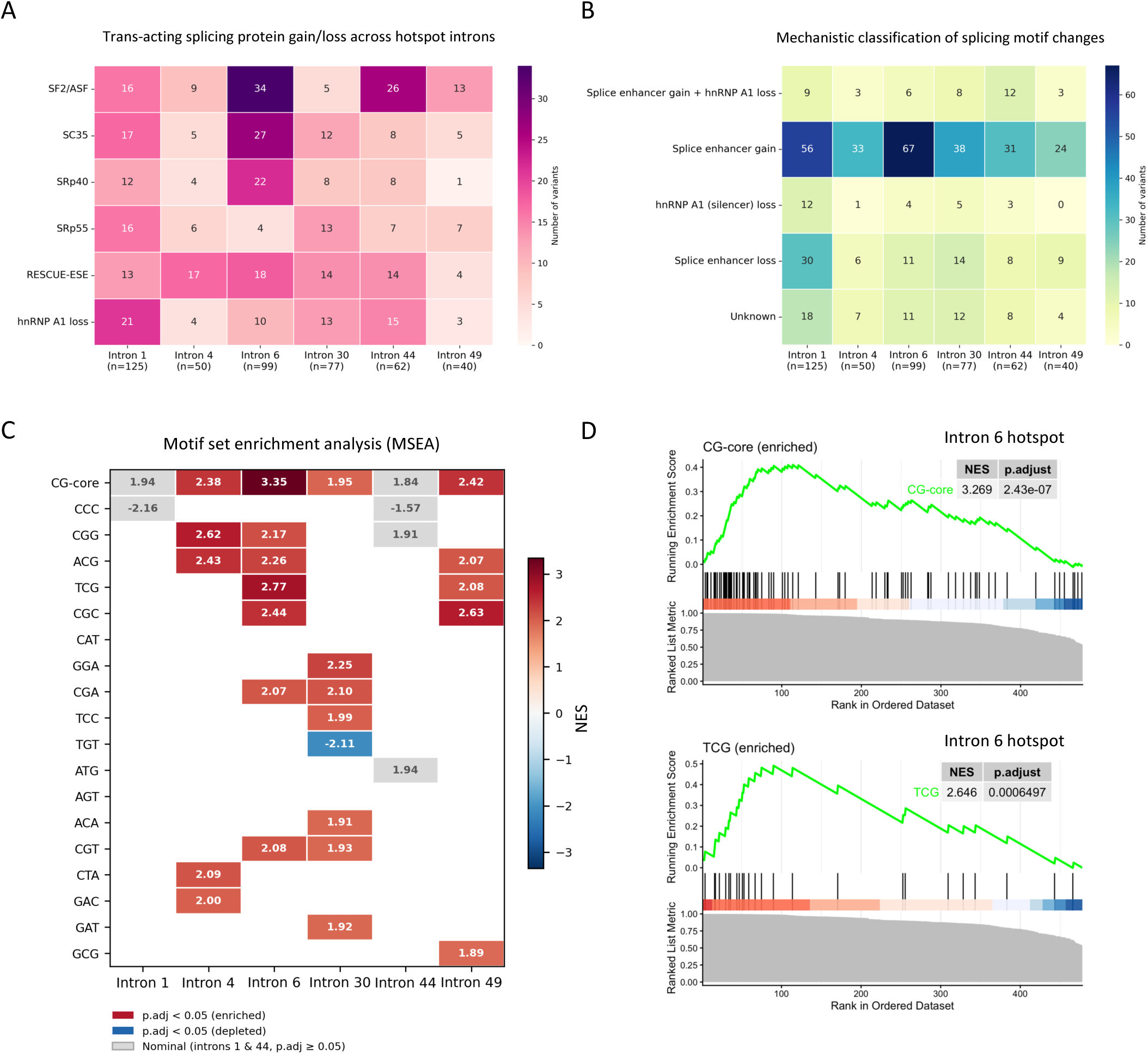
Motif disruption and motif-enrichment landscape across COL4A5 hotspot introns. (A) Heatmap showing the number of H/M impact spliceogenic variants per intron predicted to gain a splicing enhancer motif for each individual trans-acting factor, or to lose the hnRNP A1 silencer motif based on Alamut Visual Plus predictions (ESEfinder, and RESCUE-ESE). (B) Heatmap summarizing the overall mechanistic classification of the same H/M impact variants derived by applying a rule-based annotation to the ESEfinder and RESCUE-ESE predictions: variants were called “Splice enhancer gain” if they created or strengthened any enhancer motif (SF2/ASF, SC35, SRp40, SRp55, or RESCUE-ESE), “hnRNP A1 (silencer) loss” if they abolished an hnRNP A1 binding site, “Splice enhancer gain + hnRNP A1 loss” if both occurred concurrently, “Splice enhancer loss” if only an enhancer-loss event was detected with no concurrent gain, and “Unknown” if no motif change was predicted. When a variant showed both an enhancer gain and an enhancer loss (at the same or different factors), the gain call took precedence. (C) MSEA summary heatmap shows normalized enrichment scores (NES) across hotspot introns. Colored cells show FDR-significant enriched (red) and depleted (blue) motifs. Light gray cells show non-significant nominal enrichment/depletion to highlight the trends that do not pass the FDR. Number are NES values. (D) Example enrichment plots for two significant motifs from intron 6 hotspot. Representative MSEA enrichment curves for top enriched motif sets (CG-core and TCG). Insets report NES and adjusted p-value.

We next asked whether the mutant sequences created by these spliceogenic substitutions were enriched for specific motifs. MSEA of ranked mutant 7-mers showed recurrent enrichment of CG-containing motifs, especially CG-core, in introns 4, 6, 30, and 49 (**Fig. 6C**). Intron 6 showed the strongest CG-core (NES = 3.269, p.adj = 2.43 × 10⁻7) and TCG set signal (NES = 2.64, p.adj =0.000649) (**Fig. 6D**). In introns 1 and 44, CG-core also showed a positive trend but did not reach False discovery rate (FDR) significance and is shown in gray. Together, these results suggest a shared mechanism across *COL4A5* intron hotspots: spliceogenic variants frequently create CG-rich motifs that potentially serve as binding sites for trans-acting enhancer proteins, with the strongest statistical support for this CG-core enrichment observed in intron 6 (**Table S12**).

## Discussion and conclusion

Non-coding variants remain the largest unexplored source of missing heritability in Mendelian disease. Deep-intronic variants that create pseudoexons are now recognized as a recurrent cause of otherwise unsolved cases across a wide range of disorders all converging on a single cryptic splice donor site and all amenable to correction by a single splice-switching ASO. However, with growing clinical use of WGS, the potential to detect such variants is increasing, however their recognition as disease-causing is impaired by the limitations of *in silico* prediction tools. Here we tested the hypothesis that pathogenic deep-intronic variants are not uniformly distributed but converge on a small number of mechanistically vulnerable “hotspots,” where many independent sequence changes are funneled through the same aberrant splicing event. Using *COL4A5* and X-linked Alport syndrome as a tractable model, we show that such hotspots exist, can be predicted *ab initio* from sequence, and can be neutralized with a single antisense oligonucleotide irrespective of the underlying variant.

The first key finding of this study is the recurrence of pathogenic deep-intronic variants within a narrow 160bp region of intron 6 of the *COL4A5*. Identification of the same variant in multiple independent families, together with several additional variants converging on two splicing outcomes (42bp and 147bp pseudoexon retention), strongly argues against a founder effect and instead supports a genuine, mechanistically driven mutational hotspot – meaning that variants occuring in this region are more likely to be spliceogenic. This distinguishes our finding from the classical paradigm of a single recurrent deep-intronic founder allele, exemplified by the splice-mediated insertion of an Alu sequence in *COL4A3* in patients from Réunion Island (23) or the *CEP290* c.2991+1655A>G variant, which alone accounts for up to 10% of *CEP290*-related Leber congenital amaurosis (LCA10) in European and North American populations (24, 25). Rather than one ancestral variant propagated through a population, the *COL4A5* intron 6 hotspot is characterized by numerous independently arising, molecularly distinct substitutions that converge on a shared splicing mechanism - a pattern more consistent with an inherently vulnerable cis-regulatory architecture than with descent from a common ancestor.

The second key finding is the unbiased discovery of several deep-intronic hotspots using saturation ISM. This analysis identified several introns with significant positional distribution of high and moderate impact spliceogenic variants. Notably, intron 6, specifically the region of our interest, was identified as a potential hotspot. This analysis revealed that approximately 21% of all possible single-nucleotide substitutions (99 of 480) in intron 6 hotspot were predicted to have high or moderate impact on splicing. In addition to intron 6, we also identified hotspot regions in introns 1, 4, 30, 44 and 49. In intron 4 hotspot (chrX:108568011-108568117 hg38), we and others (4, 26) have identified variants in patients with Alport syndrome, providing further validation of our hypothesis and approach. Intron 44 was another interesting finding as we already identified one patient with a variant in this region. Further, retrieval of our database specified for this variants region, led to the identification of 6 additional patients with variants in this region among them two with high AlphaGenome score. Unfortunately, we did not have access to patient cells to verify the impact of these variants thus remaining unclassified. This will compel us to validate the findings with orthogonal methods. These variants consequently remain of uncertain significance, but the concordance between clinical ascertainment and unbiased *in silico* prediction supports intron 44 as the next priority target for future functional validation.

The MSEA of all hotspot introns provided mechanistic insight into the sequence features underlying splicing vulnerability in this region. The strong enrichment of CG-containing motifs (CG-core, TCG, CGC, ACG, CGG, CGA and CGT) among the top-ranked spliceogenic substitutions suggest that the creation of CpG-dinucleotide containing sequences is a key driver of pseudoexon activation. Following Gao *et al.* (27), CG dinucleotides can function as intronic splicing enhancers (CG-core ISEs), whereas the TAGG motif recognized by hnRNP A1/A2 acts as a silencer. Our enrichment of CG-core 7-mers among H/M-impact variants is therefore consistent with spliceogenic substitutions that either disrupt TAGG silencer elements or create CG-containing sequences. Notably, the intron 6 hotspot region is CpG-poor and any new change in CpG could change in local CpG density, making it more likely to shift the chromatin state. There is well established evidence that DNA methylation regulated mRNA splicing by at least two mechanisms, first by modulating the elongation rate of RNA polymerase II by CTCF and methyl-CpG binding protein MeCP2, and second by binding to the heterochromatin protein I (HP1) that recruits splicing factors (28–32).

AlphaGenome represents an advance over earlier splice prediction tools in its ability to model complex regulatory effects beyond canonical splice site motifs. However, several experimentally confirmed pathogenic variants in our dataset were scored as “No Impact” by AlphaGenome, underscoring a persistent gap between computational prediction and biological reality. These *in silico* predictions should therefore be interpreted alongside functional validation, particularly assays performed in patient-derived cells as it captures the native splicing landscape in its full complexity. As sequence-to-function models continue to be trained on larger and more diverse splicing datasets, we anticipate that their sensitivity to deep intronic and regulatory-motif-driven variants will continue to improve, progressively narrowing this gap and further strengthening the role of computational prediction in variant prioritization.

The identification of recurrent deep intronic mutational hotspots with well-defined splicing consequences opens an ASO-mediated therapeutic avenue. In the context of *COL4A5* intron 6 hotspot, the pseudoexon inclusion events identified in this study represent ideal targets for single ASO-mediated correction. Both the 147 or 42bp pseudoexon inclusion events use the same strong cryptic splice donor site (c.385-618), such that masking this site restored the normal splicing regardless of which upstream variant activates it.

The ASO used in this study was previously developed and shown to be effective in fibroblasts derived from a patient (P16) carrying the 42bp retention variant (6). We extended this approach by testing the same ASO in URECs derived from a patient carrying a distinct variant (c.385-707G>T) producing the 147 pseudoexon, and confirmed splicing correction at both the mRNA and protein level, demonstrating that a single ASO can rescue variants generating either aberrant transcript. This “one ASO, multiple variants” paradigm significantly enhances the feasibility and clinical translatability of this therapeutic approach. In vitro proof-of-concept studies in patient-derived cells and kidney organoid models (6, 7) will further strengthen the robustness of such therapies as they move towards clinical development.

In summary, this study identified deep intronic mutational hotspots using unsupervised *in silico* saturated mutagenesis in *COL4A5* introns, affecting a substantial number of patients across diverse ethnic and geographic backgrounds. Our findings underscore three key principles with broad applicability beyond Alport syndrome. First, deep intronic hotspots should be identified and systematically screened in genetically unsolved cohorts, second, functional validation of splicing variants requires complementary experimental approaches in parallel to *in silico* prediction and third, the precise characterization of pseudo-exon activation mechanisms opens opportunities for ASO-based therapeutic interventions. As genome sequencing becomes increasingly integrated into clinical diagnostics, the systematic identification and functional characterization of deep intronic variants will be essential to close the diagnostic gap in Mendelian diseases and to unlock new avenues for precision therapy.

## Methods and Materials

### Patient and Ethical Approval

All patients provided written informed consent to clinical and scientific procedures. The studies were approved by ethics committees: Comité de Protection des Personnes pour la recherche biomédicale Ile de France and the Friedrich-Alexander University Erlangen-Nürnberg (approval number: 251_18 B) and by Technical University of Munich.

### Call for variations in the region hg38 ChrX:108570700-108570800

Our gene capture panel has been described in Boisson et al. 2023(4). This diagnostic panel is regularly updated based on variants described in the literature. Since 2018, it includes a capture probe in intron 6 of *COL4A5* at position (chrX:108570700-108570800) after the identification of a pathogenic variant by other group (17). In total, 2,680 patients with suspected glomerular nephropathy were sequenced between January 2018 and March 2026 at Necker enfants malades Hospital. In this work, a retrospective variant calling was performed for variants located in this captured region (Genome version: GRCh38, Transcript: NM_33380.3).

### AlphaGenome-based analysis of variants and *in silico* mutagenesis across all introns of *COL4A5*

We first checked the impact of deep intronic variants on splicing using SpliceAI lookup portal (August 7th, 2026) from broad institut (https://spliceailookup.broadinstitute.org/). Next, the AlphaGenome(19) (v0.5.1) sequence-to-function model was accessed via API for variant effect predictions, using splice_junction and splice_site_usage modalities. We performed *in silico* mutagenesis analysis on all 52 introns of the *COL4A5* gene by extracting intronic sequences and making a dictionary of three possible nucleotide substitutions for each position and running AlphaGenome using an in-house script. We used AlphaGenome’s empirical quantile scores which rank each variant against a background distribution derived from common variants (MAF>0.01 in any gnomAD v3 population), to classify each variant’s predicted impact on splicing based on quantile thresholds applied to the splice-site usage score (High impact, ≥ 0.999 [top 0.1%]; Moderate impact, ≥ 0.99 [top 1%]; Low impact, ≥ 0.95 [top 5%]); variants below top 5% threshold were labelled “No impact”. To first test whether high- and moderate-impact variants were non-uniformly distributed along each intron we used Kolmogorov-Smirnov (KS) test. This test report two values (D and pvalue) with larger D and smaller pvalue means that these variants are not evenly spread in the intron. Then localized enriched intervals were identified with sliding-window enrichment analysis with FDR control.

After discovering deep intronic hotspots, we performed motif-set enrichment analysis (MSEA) on ISM substitutions, using mutant 7-mers scored by their AlphaGenome splice-site usage quantile as input for a pre-ranked MSEA against motif sets composed of 7-mers sharing a common nucleotide core. ISS and ISE motifs within each 7-mer were mapped using ESEfinder 3.0 (33, 34), RESCUE-ESE (35) and Alamut Visual Plus version v1.13.

The script used to automate mutagenesis, genomic track export, variant impact prediction score generation, and motif enrichment analysis is available on GitHub (https://github.com/hassansaei/Saei_COL4A5_Intron6_hotspot). To visualize these predictions, we developed multiple Python notebooks (available in the repository) that receive variant impact prediction scores as input and generate the visualizations presented in this study. Results from AlphaGenome analysis on all introns of *COL4A5* gene are deposited on Zenodo (DOI: 10.5281/zenodo.19854065).

### Minigene splicing assay

The pSPL3 minigene splicing assay was used to characterize the splicing impact of variants in the hotspot region. We first generated a wild-type pSPL3 construct containing *COL4A5* exon 5 (45-bp), exon 6 (63-bp), and exon 7 (54-bp) with flanking intronic sequences (∼80-100bp), along with the hotspot region and its flanking sequences between exons 6 and 7, using the Takara SNAP Assembly kit (Takara Bio). This kit enabled seamless fusion of multiple overlapping PCR fragments to the pSPL3 backbone. Including flanking exonic sequences alongside the deep intronic region more accurately recapitulates the *COL4A5* splicing efficiency *in vitro*. Next, we introduced 10 intronic variants, including c.385-679T>C, c.385-714G>A, c.385-716G>A, c.385-719G>A, c.385-756C>T, c.385-660A>G, c.385-708G>A, c.385-708G>C, c.385-756C>G, and c.385-752C>G using the Q5 Site-Directed Mutagenesis kit (New England Biolabs), with variant-specific primers designed using NEBaseChanger (). All vectors were sequenced by Oxford Nanopore (ONT) whole-plasmid sequencing service at the Genomics Platform at Imagine Institute. All primers used in this study are listed in **Table S13** and the ONT plasmid sequencing results are deposited on Zenodo (DOI: 10.5281/zenodo.19854065).

### Cell culture

All reagents were purchased from Gibco unless otherwise stated. HEK293T cells were cultured in Dulbecco’s Modified Eagle’s Medium (DMEM) high glucose supplemented with 10% fetal bovine serum (FBS), 2 mM L-glutamine, 100 U/ml penicillin, and 100 μg/ml streptomycin at 37°C in a humidified 5% CO₂ incubator, maintained at 80-90% confluency and passaged at a 1:5 ratio using 0.25% trypsin-EDTA. Cells were transfected with pSPL3 vectors using Lipofectamine 2000 (Thermo Fisher Scientific) to study aberrant splicing induced by deep intronic variants in intron 6.

Human urine-derived immortalized epithelial cells (URECs) from both AS 013 (carrier mother) and AS 015 (affected son with X-linked AS) were generated in Erlangen as described by Zhou et al. (36) and immortalized by lentiviral transduction with a vector expressing SV40 large T antigen with a puromycin-resistance gene. They were maintained in Renal Epithelial Cell Growth Basal Medium (Lonza Bioscience) supplemented with 2μg/ml puromycin (for immortalization-mediated resistance) and 1% penicillin-streptomycin at 37°C in 5% CO₂; these patient-derived cells were used to assess *COL4A5* mRNA and protein expression after ASO treatment.

### ASO transfection in URECs

The functional 2’O-Me full-Phosphorothioate (PS, indicated by *) ASO with sequence [U*U*C*A*A*A*U*A*C*C*U*G*U*A*U*G*C*C*A*C] and scrambled ASO with sequence [A*G*A*C*G*C*A*C*U*A*A*U*A*A*G*C*A*U*G*C*A] used in this study have been previously tested on patient-derived cells (P16)(6). This ASO was designed to mask the cryptic splice donor site at intron 6 (CAG|GTA; c.385-617/618). Patient AS 015-derived URECs were seeded at 400,000 cells per well in 6-well plates. After 24 hours, the medium was replaced with Opti-MEM (Thermo Fisher Scientific). The ASO-Lipofectamine complex was prepared by diluting 5μl Lipofectamine 3000 in 245μl Opti-MEM (tube 1) and 2μl of 100μM ASO in 250μl Opti-MEM (tube 2). The solutions were combined, mixed gently, and incubated for 15 minutes at room temperature before being added dropwise to each well. Negative control and untreated wells received equivalent volumes of transfection medium. After 6 hours of incubation, the transfection medium was removed and replaced with a complete growth medium (without any antibiotics for URECs). Cells were cultured for 48 hours for RNA and protein analysis.

### Total RNA extraction, RT-PCR and RT-qPCR

Total RNA was extracted from HEK293T cells or URECs using the Qiagen RNeasy Mini Kit (Qiagen) according to the manufacturer’s protocol. First-strand cDNA was synthesized using SuperScript II (thermo Fisher Scientific) after treating samples with RQ1 RNase-free DNase I (Promega Corporation). The resulting cDNA was used for RT-PCR with target-specific primers (**Table S1**). PCR products were separated on a 1.5% agarose gel. *COL4A5* gene expression was analyzed by qPCR using a single primer pair that amplifies both wild-type and mutant transcripts. qPCR was performed using iTaq Universal SYBR Green Supermix (Bio-Rad) on a Bio-Rad CFX Opus 384 system.

### Fragment analysis

To quantify differences in the relative abundance of wild-type and mutant *COL4A5* transcripts, we performed fragment analysis using a FAM-labeled forward primer. Using qPCR, we determined the number of cycles required for diluted cDNA (1:10) to reach the exponential phase of amplification; based on this, we used 32 cycles to avoid amplification saturation prior to fragment analysis. For capillary electrophoresis, 0.5µL of GeneScan 600 LIZ DNA size standard (Thermo Fisher Scientific) was added to 18.5µL of formamide (Sigma-Aldrich), and 1µL of the PCR product was added to this mixture. Samples were heated at 95°C for 5 minutes. Data were analyzed using the Thermo Fisher Cloud service (PeakScanner software).

### Targeted RNA sequencing

We performed *COL4A5* targeted RNA sequencing on patient-derived cells as we described previously(4). In short, first strand cDNA was synthesized using SuperScript IV VILO Master Mix (Thermo Fisher Scientific) and the synthesis of second strand was performed using Second Strand cDNA Synthesis Kit (Thermo Fisher Scientific). The double-stranded cDNAs were purified using AMPure XP Reagent (Beckman Coulter), and 50ng of cDNA was used to prepare the next-generation sequencing targeted RNA-seq libraries using a Twist technology. Paired-end sequencing (150 bp) was performed on AVITI (Element Bioscience). The FASTQ files were aligned to the GRCh37 assembly of the human genome using Illumina Dragen software, and the visualization of the junctional reads with Sashimi plots was performed using the ggsashimi(37). The BAM files are desposited on Zenodo (DOI: 10.5281/zenodo.19854065).

### Western Blot analysis of a5(IV) collagen expression

Whole-cell extract of URECs transfected with ASO or control scramble ASO, were performed as previously described (38). In short, cells were sonicated for 5 seconds at 50% amplitude (Bandelin Sonoplus HD 2070, Bandelin electronic, Berlin, Germany) in an extraction buffer (8 M urea, 10% glycerol, 1% SDS, 10 mM Tris-HCl pH 6.8) supplemented with cOmplete protease inhibitors (Roche, Mannheim, Germany). Protein concentrations were determined using the DC Protein Assay (Bio-Rad, CA) following the manufacturer’s protocol. Proteins were resolved by SDS-PAGE and transferred onto PVDF membranes (Millipore, Bedford, MA, USA) at 115 mA per membrane for 2 h in a Mini Trans-Blot Cell system (Bio-Rad, Hercules, CA, USA). Membranes were blocked for 1 hour at room temperature in 5% milk in TBS-T, then incubated overnight at 4°C with a primary antibody against collagen α5 (IV)(clone H-53, Chondrex (Redmond, WA, USA), 1:500). An HRP-conjugated secondary antibody (Goat tanti-rat HRP, Jackson ImmunoResearch (Cambridgeshire, UK), 1:2000) was applied for 30 minutes at room temperature. Membranes were washed 3 × 5 minutes with TBS-T between incubations, and after secondary antibody incubation were washed 4 × 10 minutes with TBS-T followed by a 2-minute rinse in phosphate-buffered saline. Signals were detected using an ECL system (GE Healthcare, Munich, Germany).

### Statistical analysis and data visualization

All statistical analyses were performed using GraphPad Prism version 11. AlphaGenome analyses (cohort variants, and single-nucleotide substitution scoring across all introns of the *COL4A5*) were run in Python (Jupyter notebooks, Python 3.11) using the alphagenome-env conda environment specified in the companion repository (https://github.com/hassansaei/Saei_COL4A5_Intron6_hotspot); quantile-based splice-impact summaries, heatmaps, and related figures were generated in the same environment. Motif-context construction, preranked motif-set enrichment inputs, and associated Python outputs were produced with alphagenome-env; complementary MSEA panels and figures were generated in R with the r-motif-gsea-env environment, as documented in that repository. Welch’s t-test was applied to compare healthy control and AS015 samples, and one-way ANOVA was used for ASO treatment analyses. The Mann–Whitney U test was used to assess relative signal intensity in Western blot experiments, and the resulting data were presented as bar graphs (mean ± SEM). Two-way ANOVA was used for sample comparisons in fragment analysis.

## Data Availability

All data produced are available online at DOI: https://doi.org/10.5281/zenodo.19854065

## Acknowledgments

We would like to first thank all patients and their families for supporting this work. We sincerely thank the Google DeepMind research team involved in generating the AlphaGenome model, especially Natasha Latysheva, for providing us with special access with an increased API quota, which enabled us to perform large-scale predictions. G.D received support from ORKID. K.M.R. was supported by a grant from the German Research Foundation (DFG; Project No.: 519309154).

Data from the National Genomic Research Library (NGRL) used in this research are available within the secure Genomics England Research Environment. Access to NGRL data is restricted to adhere to consent requirements and protect participant privacy. Data used in this research include: Genomic variants pulled from the interactive variant analysis store for patients sequenced in the 100,000 genomes project linked by their participant ID to basic clinical information via the participant explorer. Relevant findings were exported under project ID 1479. Access to NGRL data is provided to approved researchers who are members of the Genomics England Research Network, subject to institutional access agreements and research project approval under participant-led governance. For more information on data access, visit: https://www.genomicsengland.co.uk/research

## Author Contributions

H.S. designed and performed research and analyzed data; G.D. supervised research; B.A., N.K., M.O., O.G., and V.M. performed research; F.J.W., K.M.R., C.G., O.Gr., C.P., L.L., M.R., and C.B. contributed clinical data and patient information; M.S.W. provided patient-derived cells and edited the paper; and H.S., C.A and G.D. wrote and edited the paper.

## Declaration of Generative AI and AI-assisted technologies in the manuscript preparation process

During the preparation of this work the authors used Anthropic Claude (Opus 5), and Cursor in order to assist with data analysis scripting, figure generation, and to support language refinement and clarity of expression in the manuscript. After using these tools, the authors reviewed and edited the content as needed and take full responsibility for the content of the published article

