## Supplemental Tables and Figures for "Mapping intronic mutational hotspots by *in silico* mutagenesis enables single antisense oligonucleotide correction of multiple variants"

**This PDF file includes:**

Figures **S1 to S5**

Tables **S1 to S13**

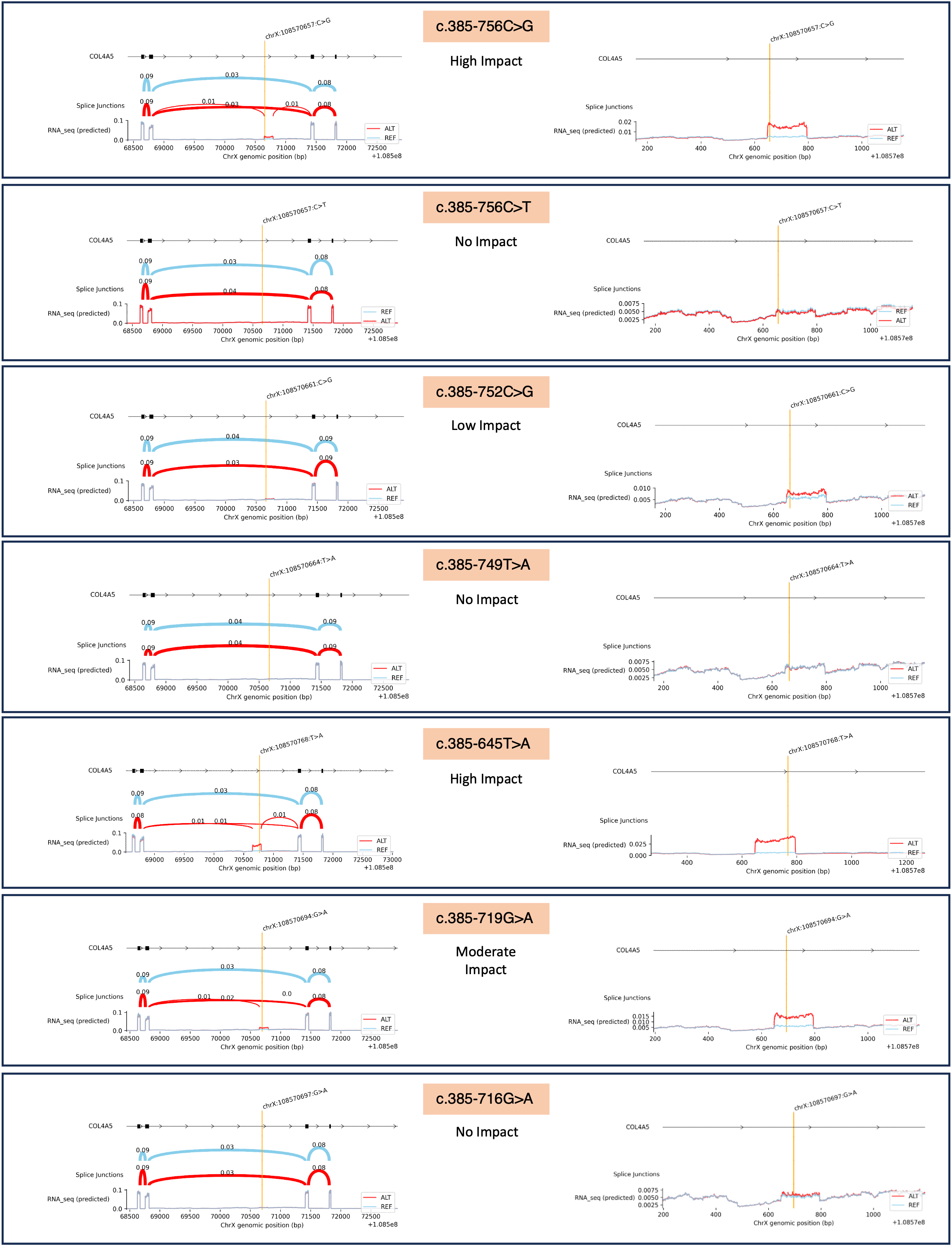

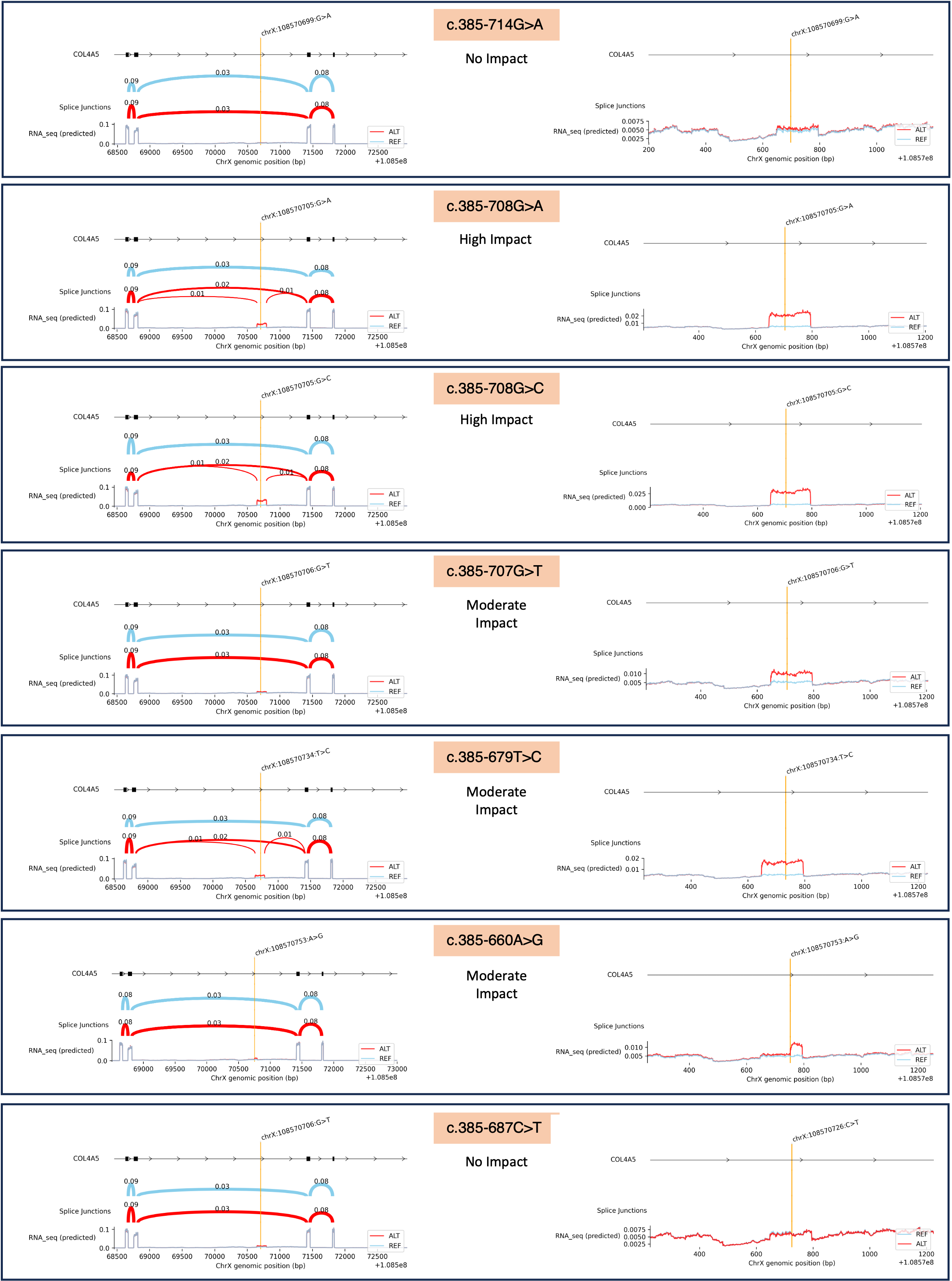

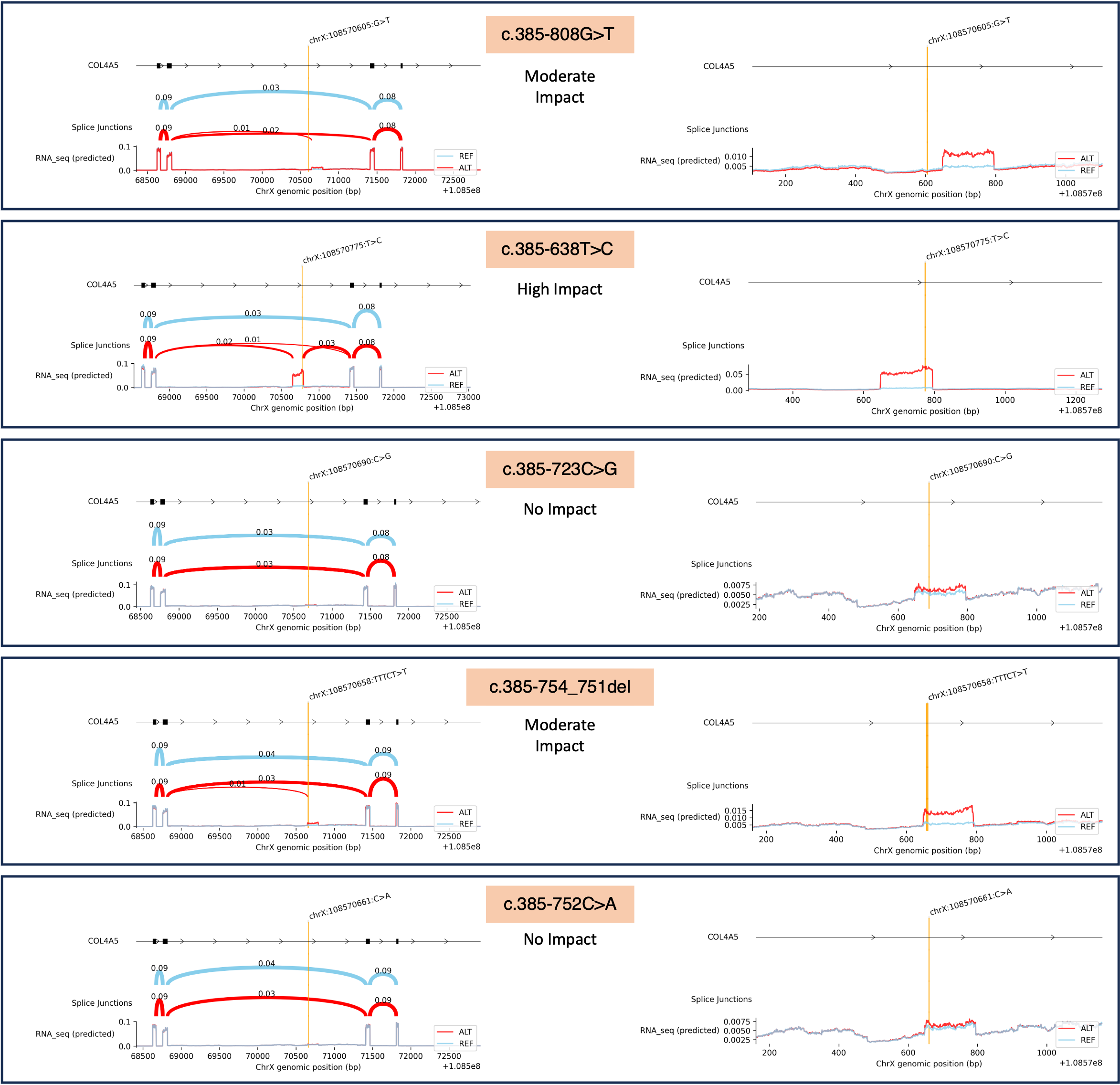

**Fig. S1. AlphaGenome splicing analysis of 19 deep intronic variants in *COL4A5* with their predicted Sashimi plots.** Comparative splice prediction plots generated by AlphaGenome show the effects of each variant in the intron 6 hotspot on predicted exon-intron junction usage and transcript coverage in kidney tissue. Each pair of panels shows the reference (REF) and alternative (ALT) allele splicing profiles for an individual variant (variants indicated in orange). The left plot in each panel displays predicted splice-junction strength and isoform models derived from tissue-specific RNA-seq data. The right plot shows the zoomed-in version of the predicted difference in splice junction probability between reference (skyblue) and altered (red) alleles. Variants with strong pseudoexon activation potential have higher RNA-seq predicted score.

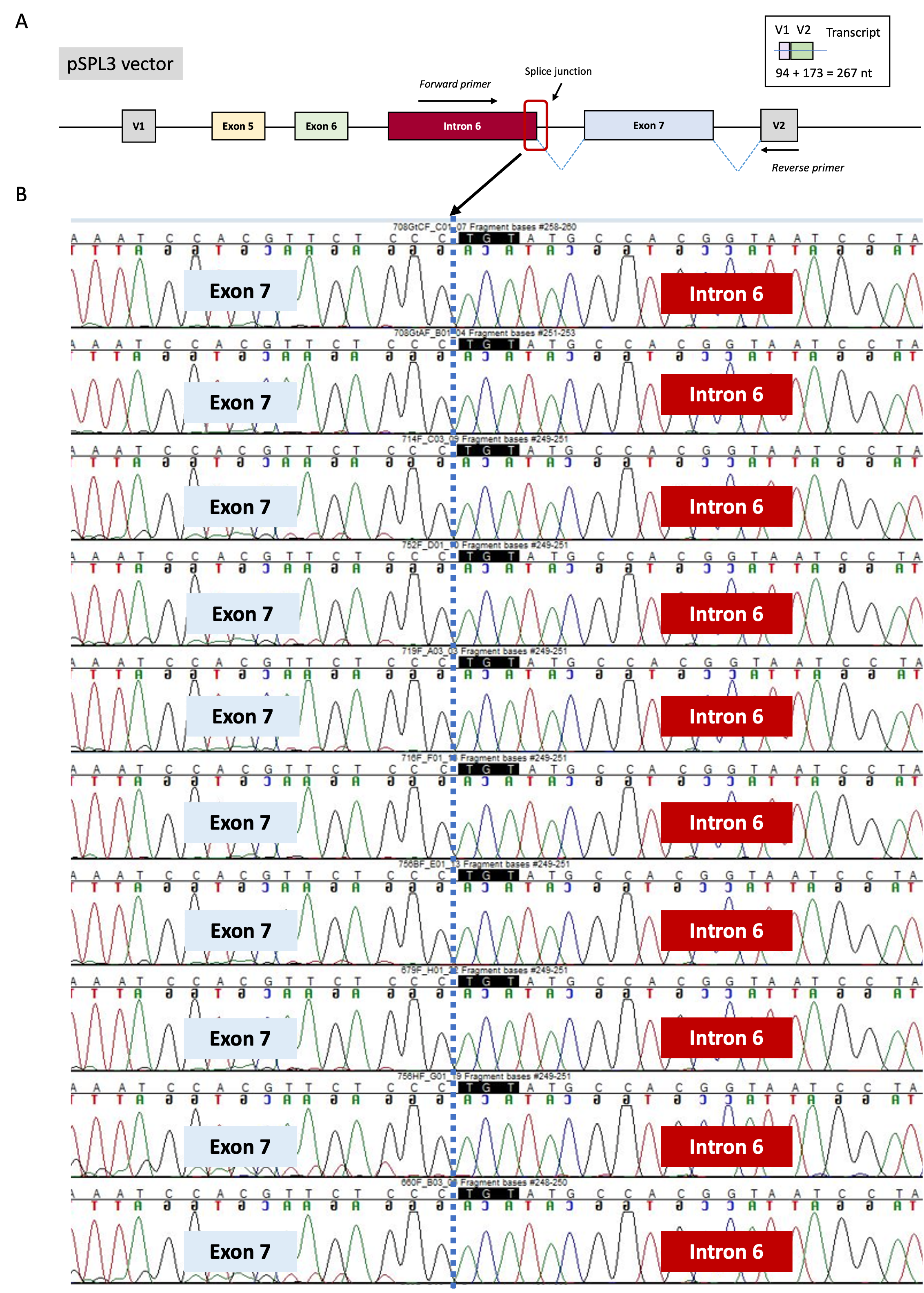

**Fig. S2. Sequencing pSPL3 encoded mRNAs in HEK293 cells harboring 10 deep intronic variants in intron 6.** (A) All cDNAs were amplified by PCR using an internal forward primer bound to the pseudoexon in intron 6 and a reverse primer bound to V2 exon in the pSPL3 vector.

(B) The PCR products were sequenced by the Sanger method and analyzed using the Sequencher software. The intron 6-exon 7 aberrant junction is highlighted by blue dashed line.

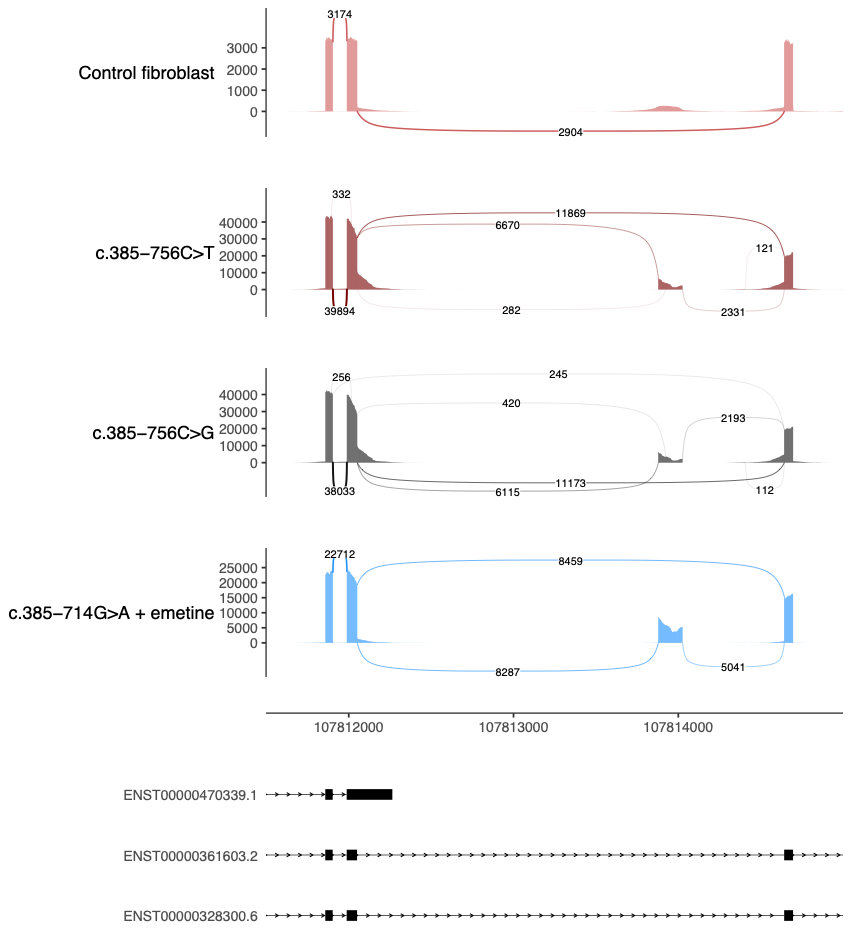

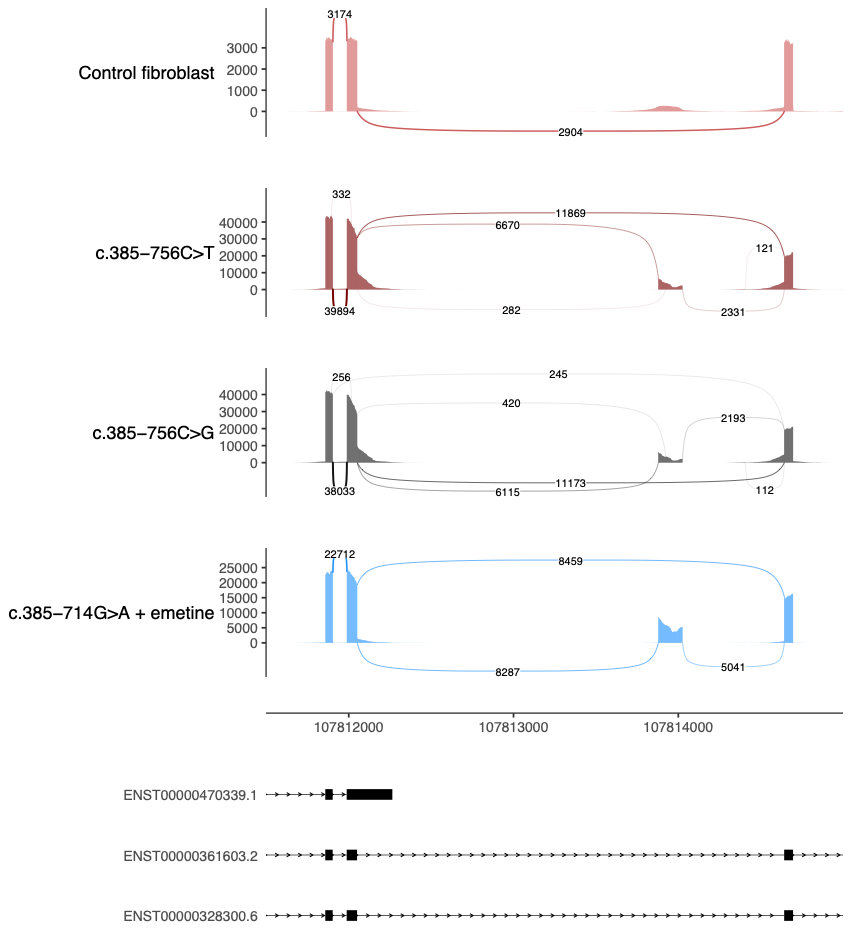

**Fig. S3. Targeted RNA sequencing on patient cells confirmed the pathogenicity of deep intronic variants with weak minigene results.** Sashimi plots for control fibroblasts and variants; c.385-756C>T, c.385-756C>G without NMD inhibitor (emetine) and c.385-714G>A with emetine. The plots were generated by ggsashimi python package with the minimum number of reads supporting a junction to be displayed set to 100.

**Fig. S4.**
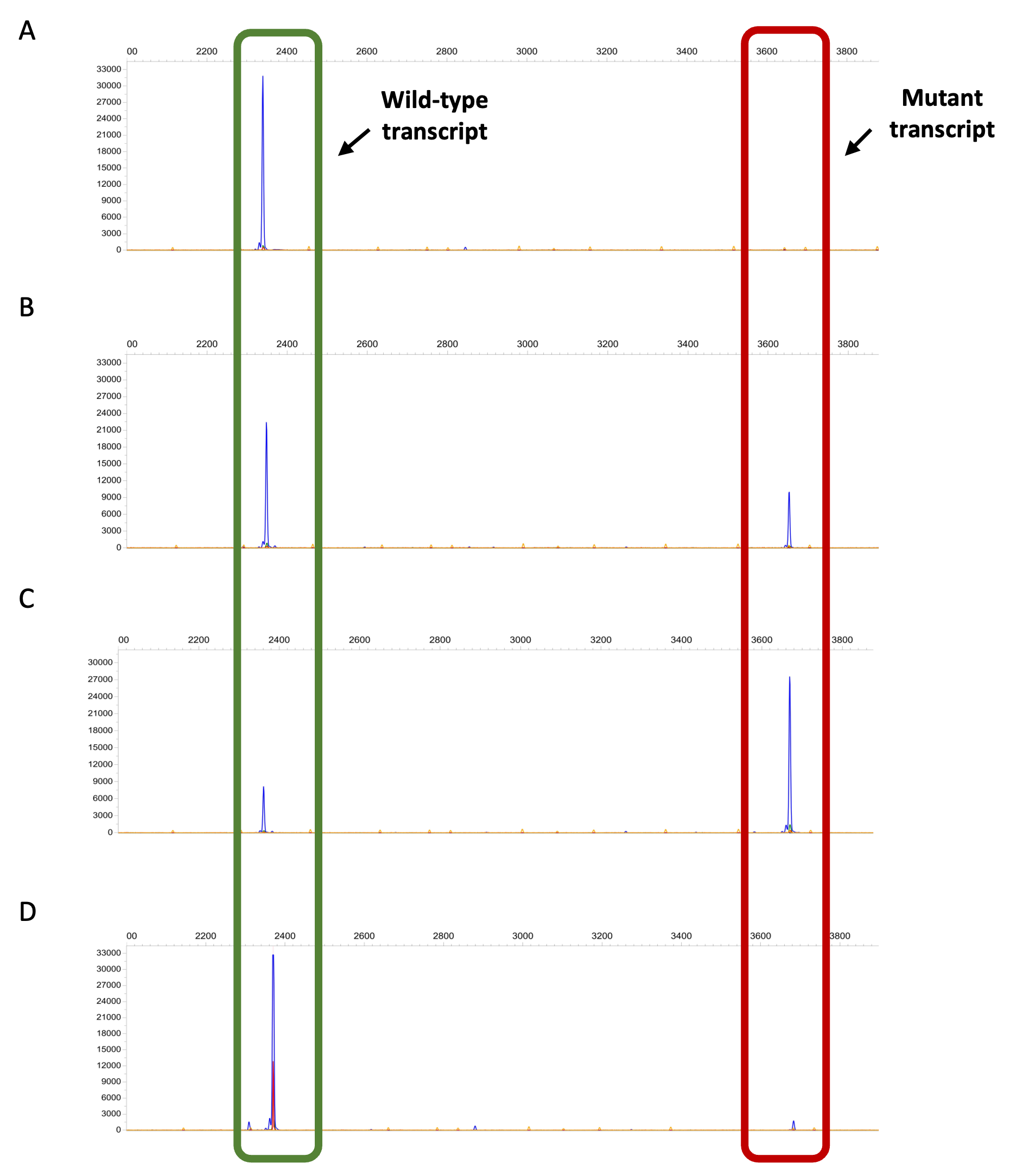
 **Fragment analysis of wild-type and mutant *COL4A5* transcripts in URECs with and without ASO treatment.** (A) Control URECs show only the short wild-type transcript fragment. (B) URECs from the AS 015 Alport patient without NMD inhibition display a shorter wild-type peak and the appearance of a mutant transcript fragment. (C) In Alport patient URECs treated with an NMD inhibitor, the mutant transcript becomes the predominant peak. (D) AS 015 patient URECs treated with ASO without NMD inhibition show successful restoration of the wild-type transcript. The fragment results were analyzed by Peak Scanner software (Thermo Fisher cloud service).

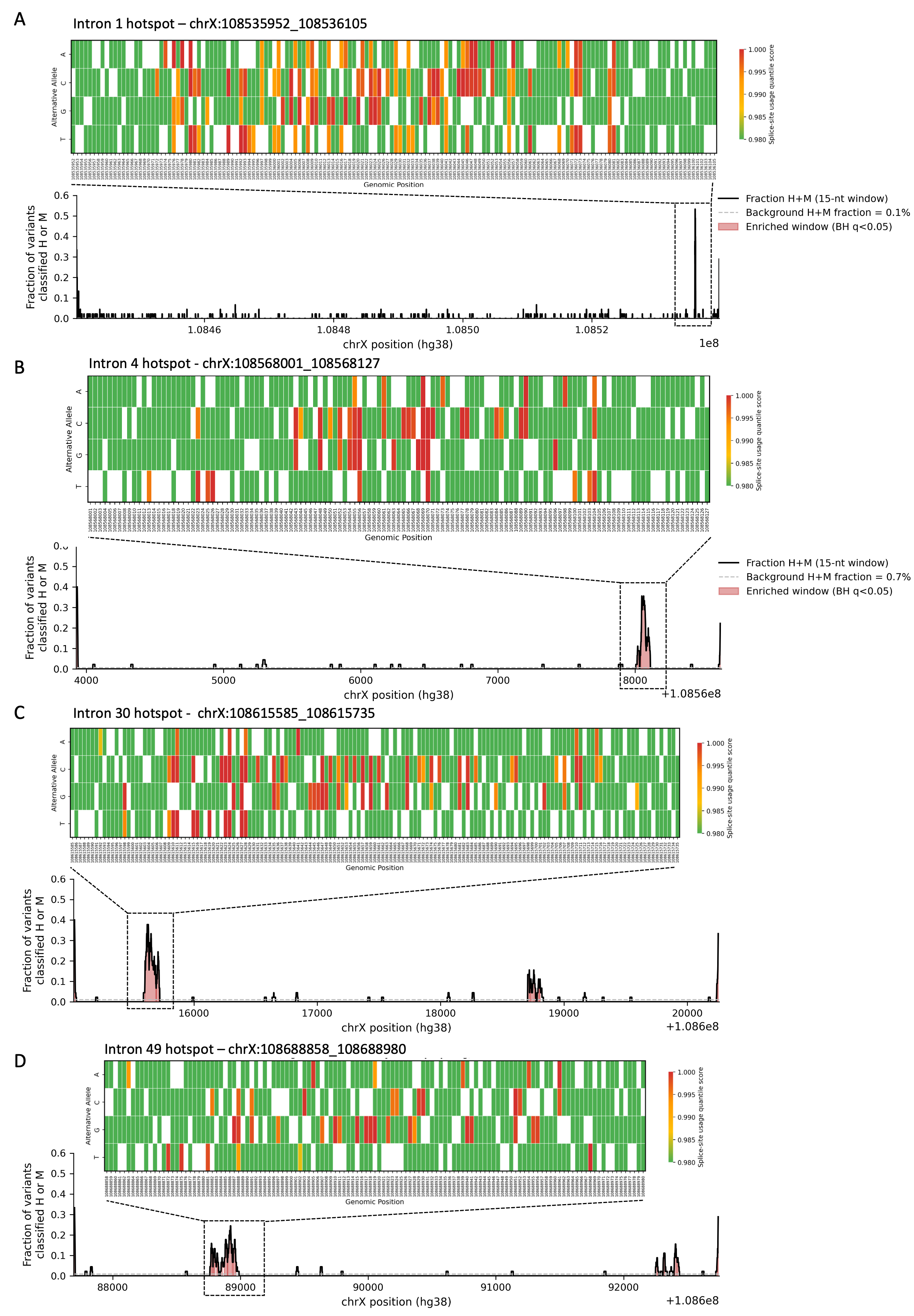

**Fig. S5. Spatial hotspot profiles in COL4A5 intron 1, 4, 30 and 49.** (A) ISM landscape across the deep intronic hotspot in intron 1. Each column corresponds to a single genomic position and each row to one of the three possible substitutions. Cells are colored by the AlphaGenome splice site usage quantile score (color scale 0.98 – 1.00; higher = more disruptive to splicing); white cells indicate the reference allele at that position. Variants predicted to most strongly disrupt splicing appear in red. (B-D) ISM landscape across the deep intronic hotspot in intron 4, 30 and 49.

**Table S1.** Results of AlphaGenome splicing scoring of 19 deep intronic variants identified in the intron 6 hotspot

| Variation | Genomic Coordinate (Hg38) | Change | Impact Classification | Max Quantile Score | Max Raw Score | Mean Quantile Score | Splice Junction Quantile | Splice Site Quantile* |
| --- | --- | --- | --- | --- | --- | --- | --- | --- |
| c.385-645T>A | 108570768 | T>A | High Impact | 0.999982 | 2.996094 | 0.999895 | 0.999982 | 0.999846 |
| c.385-708G>C | 108570705 | G>C | High Impact | 0.999978 | 2.810547 | 0.999901 | 0.999978 | 0.999817 |
| c.385-708G>A | 108570705 | G>A | High Impact | 0.99996 | 2.304688 | 0.999789 | 0.99996 | 0.999558 |
| c.385-756C>G | 108570657 | C>G | High Impact | 0.999956 | 2.236328 | 0.998197 | 0.999956 | 0.999459 |
| c.385-679T>C | 108570734 | T>C | High Impact | 0.999946 | 2.091797 | 0.999615 | 0.999946 | 0.99916 |
| c.385-719G>A | 108570694 | G>A | Moderate Impact | 0.999874 | 1.50293 | 0.999384 | 0.999874 | 0.998696 |
| c.385-660A>G | 108570753 | A>G | Moderate Impact | 0.999817 | 1.276367 | 0.997871 | 0.999817 | 0.995289 |
| c.385-707G>T | 108570706 | G>T | Moderate Impact | 0.999804 | 1.248047 | 0.998193 | 0.999804 | 0.995884 |
| c.385-752C>G | 108570661 | C>G | Low Impact | 0.999477 | 0.744629 | 0.967292 | 0.999477 | 0.968384 |
| c.385-714G>A | 108570699 | G>A | No Impact | 0.998506 | 0.363281 | 0.744411 | 0.998506 | 0.632299 |
| c.385-716G>A | 108570697 | G>A | No Impact | 0.997063 | 0.214966 | 0.912691 | 0.997063 | 0.960258 |
| c.385-756C>T | 108570657 | C>T | No Impact | 0.995289 | 0.138672 | 0.810699 | 0.995289 | 0.909199 |
| c.385-749T>A | 108570664 | T>A | No Impact | 0.990451 | 0.06189 | 0.775819 | 0.990451 | 0.805808 |
| c.385-638T>C | 108570775 | T>C | High Impact | 0.99999 | 3.628906 | 0.99904 | 0.99999 | 0.999942 |
| c.385-808G>T | 108570605 | G>T | Moderate Impact | 0.999922 | 1.819336 | 0.999383 | 0.999922 | 0.998604 |
| c.385-723C>G | 108570690 | C>G | No Impact | 0.998651 | 0.395996 | 0.725286 | 0.998651 | 0.183808 |
| c.385-752C>A | 108570661 | C>A | No Impact | 0.996404 | 0.176392 | 0.788364 | 0.996404 | 0.727613 |
| c.385-687C>T | 108570726 | C>T | No Impact | 0.986197 | 0.05014 | 0.772193 | 0.986197 | 0.906363 |
| c.385-754_385-751del | 108570658 | TTTCT>T | Moderate Impact | 0.999865 | 1.467773 | 0.999239 | 0.999865 | 0.998347 |

*****Impact Classification was assigned based on the Splice Site Max Quantile score for each variant, as detailed in Methods.

**Table S2.** Predicted splicing regulatory motif changes at intron 6 hotspot variants.

| Variation | Genomic coordinate (hg38) | Pathogenicity | Mutant 7-mer | Reference 7-mer | Predicted Motif Change |
| --- | --- | --- | --- | --- | --- |
| c.385-660A>G | g.108570753A>G | Proven | TCAGACC | TCAAACC | Gain: SRp40 |
| c.385-714G>A | g.108570699G>A | Proven | GGAAAGA | GGAGAGA | Gain: RESCUE-ESE hexamer |
| c.385-756C>T | g.108570657C>T | Proven | TTCTTTT | TTCCTTT | Unknown |
| c.385-756C>G | g.108570657C>G | Proven | TTCGTTT | TTCCTTT | Gain: RESCUE-ESE hexamer |
| c.385-752C>G | g.108570661C>G | Proven | TTTGTCT | TTTCTCT | Unknown |
| c.385-708G>A | g.108570705G>A | Proven | AAGAGTA | AAGGGTA | Loss: hnRNP A1 |
| c.385-708G>C | g.108570705G>C | Proven | AAGCGTA | AAGGGTA | Gain: SRp55; Loss: hnRNP A1 |
| c.385-679T>C | g.108570734T>C | Proven | CAGCAGT | CAGTAGT | Gain: SF2/ASF; Gain: SF2/ASF-IgM/BRCA1 |
| c.385-687C>T | g.108570726C>T | Suspected | ACATACA | ACACACA | Unknown |
| c.385-808G>T | g.108570605G>T | Suspected | AATTTAT | AATGTAT | Gain: SRp40; Loss: SRp55 |
| c.385-638T>C | g.108570775T>C | Suspected | TTCCAGG | TTCTAGG | Gain: SRp40 |
| c.385-723C>G | g.108570690C>G | Suspected | TAAGAAG | TAACAAG | Loss: SRp40 |
| c.385-754_385-751del | g.108570659_108570662del | Suspected | CCTCTCG | TTTCTCT | Unknown |
| c.385-752C>A | g.108570661C>A | Suspected | TTTATCT | TTTCTCT | Loss: SRp40 |
| c.385-707G>T | g.108570706G>T | Suspected | AGGTTAT | AGGGTAT | Gain: SRp40; Gain: SC35 |
| c.385-716G>A | g.108570697G>A | Proven | GTGAAGA | GTGGAGA | Gain: RESCUE-ESE hexamer |
| c.385-719G>A | g.108570694G>A | Proven | AAGATGG | AAGGTGG | Gain: RESCUE-ESE hexamer |
| c.385-645T>A | g.108570768T>A | Proven | AAGATGT | AAGTTGT | Gain: RESCUE-ESE hexamer |
| c.385-749T>A | g.108570664T>A | Proven | CTCACGG | CTCTCGG | Gain: SF2/ASF; Gain: SF2/ASF-IgM/BRCA1 |

*SF2/ASF, SF2/ASF (IgM-BRCA1), SC35, SRp40, and SRp55 are Exon Splice Silencer and Intron Splice Enhancer (ESS/ISE) motifs whose gain is predicted to promote pseudoexon inclusion. hnRNP A1 is an ISS factor; loss of its binding site is predicted to relieve repression, favoring pseudoexon inclusion. RESCUE-ESE hits are statistically enriched hexamers without assigned trans-acting specificity or directionality. “Unknown” indicates no predicted motif change.*

**Table S3.** Clinical characteristics of patients with suspected Alport syndrome, identified through international collaboration

| Variation in intron 6 | Family | Zygosity | | CKD stage | Age range | Hearing impairment | Occular symptomes | Remarks |
| --- | --- | --- | --- | --- | --- | --- | --- | --- |
| c.385-687C>T | I | | Heterozygous | 2 | 51-55 | no | no | Carries two variants:  1) COL4A3: c.4235G>A; p.Gly1412Asp  2) COL4A5: c.385-687C>T |
| c.385-808G>T | I | | Heterozygous | - | 46-50 | no | cataract | Proteinuria, hematuria, normal kidney function |
|  | II | | Heterozygous | - | 61-65 | no | no | - |
|  | III | | Heterozygous | - | 10-15 | no | no | Microhematuria, normal kidney function |
| c.385-638T>C | I | | Heterozygous | 5 | 31-35 | no | no | Biopsy findings suggestive of Alport syndrome |
| c.385-723C>G | I | | Heterozygous | AKI stage 3 | 31-35 | no | no | Acute kidney injury stage 3 |
| c.385-754_385-751del | I | | Hemizygous | - |  |  | - | - |
| c.385-752C>A | I-1 | | Heterozygous | - | 51-55 | - | Unknown | Proteinuric kidney disease |
|  | I-2 | | Heterozygous | - | 31-35 | - | Unknown | Proteinuric kidney disease |
|  | II | | Heterozygous | - | 51-55 | - | Unknown | Proteinuric kidney disease |
| c.385-707G>T | I | | Hemizygous | 1 | 6-10 | unknown | unknown | Variant pathogenecity has aleady been aproved on patient-derived urinary cells and skin and kidney biopsy |

**Table S4.** Systematic unbiased ISM analysis of all introns of *COL4A5* gene identified 6 potential intron hotspots.

| **Intron** | **hotspot_start** | | **hotspot_end** | **length_nt** | **min_BH_q** | **n_HM_interior** | **n_HM_in_hotspot (High/Moderate)** | **Fraction_total** | **n_total_in_hotspot** | **fraction_HM_in_hotspot** |
| --- | --- | --- | --- | --- | --- | --- | --- | --- | --- | --- |
| Intron 1 | | 108535962 | 108536095 | 134 | 6.17E-52 | 365 | 125 (37/88) | 0.342 | 402 | 0.311 |
| Intron 4 | | 108568011 | 108568117 | 107 | 1.73E-20 | 71 | 50 (27/23) | 0.704 | 321 | 0.156 |
| Intron 6 | | 108570641 | 108570801 | 161 | 2.65E-11 | 151 | 108 (48/60) | 0.715 | 483 | 0.224 |
| Intron 30 | | 108615595 | 108615725 | 131 | 9.80E-20 | 123 | 77 (41/36) | 0.626 | 393 | 0.196 |
| Intron 44 | | 108680178 | 108680314 | 137 | 4.73E-23 | 76 | 61 (11/50) | 0.802 | 411 | 0.148 |
| Intron 49 | | 108688868 | 108688970 | 103 | 1.20E-10 | 100 | 40 (16/24) | 0.40 | 309 | 0.129 |

min_BH_q, smallest Benjamini-Hochberg-adjusted q-value among the 15-nt sliding windows that define the hotspot; n_HM_interior, High and Moderate impact spliceogenic variant count in deep intron (>100 nt from the start and end of the intron); n_HM_in_hotspot, H/M variant count inside hotspot (inclusive coordinates); Fraction_total, equals hotspot concentration; n_total_in_hotspot, total ISM variants inside hotspot; fraction_HM_in_hotspot, H/M fraction among all substitutions from ISM.

**Table S5.** Clinical table of patients suspected to have Alport syndrome with deep intronic variants identified in intron 44 hotspot.

| Genomic Coordinate (Hg38) | Variant | Impact Classification | Spliceai | Sex | Clinical History |
| --- | --- | --- | --- | --- | --- |
| X:10868020G>A | c.3943-478G>A | No Impact | - | Female | CKD, arterial hypertension, diabetes |
| X:108680219A>T | c.3943-460A>T | No Impact | AG:0.33 | Male | Proven Alport syndrome. |
| X:108680231G>A | c.3943-448G>A | No Impact | AG:0.06 | Male | CKD at the age of 20. Single right kidney. Proteinuria suggestive of glomerular origin. |
| X:108680234T>C | c.3943-445T>C | High Impact | AG:0.44 | Male | Nephropathy of undetermined origin with end-stage renal disease (ESRD) at age 24. Bilateral anterior lenticonus |
| X:108680270T>C | c.3943-409T>C | Moderate Impact | AG:0.18 | Female | Hematuria |
| X:108680309CC>C | c.3943-369_3943-368del | No Impact | - | Female | CKD at age 39, arterial hypertension, atrophic kidneys. |

CKD; chronic kidney disease

**Table S6.** The list of substitutions from ISM analysis with high impact on intron 6 retention

| Variant ID | Change | Impact Classification | Overall Max Raw | Splice Junction Max Quantile | Splice Junction Mean Quantile | Splice Site Max Raw | Splice Site Mean Raw | Splice Site Max Quantile | Splice Site Mean Quantile |
| --- | --- | --- | --- | --- | --- | --- | --- | --- | --- |
| chrX:108570776:A>C | A>C | High Impact | 4.191406 | 0.99999 | 0.998514 | 0.59848 | 0.527405 | 0.999885 | 0.999844 |
| chrX:108570775:T>G | T>G | High Impact | 4.230469 | 0.99999 | 0.99997 | 0.777039 | 0.708862 | 0.99994 | 0.999918 |
| chrX:108570740:T>G | T>G | High Impact | 4.714844 | 0.99999 | 0.999987 | 0.356201 | 0.303513 | 0.999785 | 0.999722 |
| chrX:108570735:A>G | A>G | High Impact | 4.035156 | 0.99999 | 0.999981 | 0.251648 | 0.216194 | 0.999659 | 0.999533 |
| chrX:108570635:A>G | A>G | High Impact | 3.894531 | 0.999988 | 0.998336 | 0.593733 | 0.517562 | 0.999885 | 0.999844 |
| chrX:108570775:T>C | T>C | High Impact | 3.628906 | 0.999985 | 0.997294 | 0.589539 | 0.521362 | 0.999885 | 0.999844 |
| chrX:108570719:A>G | A>G | High Impact | 3.535156 | 0.999984 | 0.998531 | 0.526642 | 0.455551 | 0.999876 | 0.999833 |
| chrX:108570777:G>C | G>C | High Impact | 3.277344 | 0.999979 | 0.998162 | 0.357193 | 0.300095 | 0.999785 | 0.99971 |
| chrX:108570775:T>A | T>A | High Impact | 3.279297 | 0.999979 | 0.996319 | 0.530457 | 0.457733 | 0.999876 | 0.999833 |
| chrX:108570732:A>G | A>G | High Impact | 3.203125 | 0.999978 | 0.999964 | 0.128288 | 0.112419 | 0.999015 | 0.998843 |
| chrX:108570768:T>A | T>A | High Impact | 3.007812 | 0.999974 | 0.999956 | 0.314529 | 0.272652 | 0.999741 | 0.999679 |
| chrX:108570705:G>C | G>C | High Impact | 2.810547 | 0.99996 | 0.999935 | 0.272415 | 0.231972 | 0.999689 | 0.999598 |
| chrX:108570776:A>T | A>T | High Impact | 2.703125 | 0.999956 | 0.99993 | 0.250824 | 0.212791 | 0.999659 | 0.999512 |
| chrX:108570658:T>G | T>G | High Impact | 2.648438 | 0.999954 | 0.997259 | 0.230225 | 0.192726 | 0.999634 | 0.999444 |
| chrX:108570687:T>G | T>G | High Impact | 2.630859 | 0.999953 | 0.999922 | 0.340332 | 0.290268 | 0.999764 | 0.999699 |
| chrX:108570776:A>G | A>G | High Impact | 2.554688 | 0.999947 | 0.999916 | 0.202576 | 0.179062 | 0.999518 | 0.999385 |
| chrX:108570760:T>G | T>G | High Impact | 2.507812 | 0.999945 | 0.999907 | 0.176804 | 0.154907 | 0.999407 | 0.999222 |
| chrX:108570735:A>C | A>C | High Impact | 2.4375 | 0.999937 | 0.999899 | 0.213562 | 0.187408 | 0.99958 | 0.999425 |
| chrX:108570704:G>C | G>C | High Impact | 2.441406 | 0.999937 | 0.9999 | 0.208313 | 0.178589 | 0.99954 | 0.999369 |
| chrX:108570643:G>T | G>T | High Impact | 2.402344 | 0.999935 | 0.999894 | 0.250305 | 0.219482 | 0.999659 | 0.999565 |
| chrX:108570705:G>A | G>A | High Impact | 2.304688 | 0.999927 | 0.999888 | 0.147415 | 0.129601 | 0.99927 | 0.999056 |
| chrX:108570662:T>G | T>G | High Impact | 2.304688 | 0.999927 | 0.996677 | 0.215393 | 0.180862 | 0.99958 | 0.999371 |
| chrX:108570706:G>A | G>A | High Impact | 2.234375 | 0.99992 | 0.999881 | 0.141724 | 0.123016 | 0.999218 | 0.998988 |
| chrX:108570657:C>G | C>G | High Impact | 2.238281 | 0.99992 | 0.995876 | 0.128052 | 0.110321 | 0.999015 | 0.998795 |
| chrX:108570771:T>C | T>C | High Impact | 2.199219 | 0.999914 | 0.999865 | 0.189697 | 0.168091 | 0.999507 | 0.999344 |
| chrX:108570687:T>C | T>C | High Impact | 2.1875 | 0.999913 | 0.999857 | 0.209473 | 0.178452 | 0.99956 | 0.99938 |
| chrX:108570725:A>G | A>G | High Impact | 2.144531 | 0.999906 | 0.999858 | 0.15242 | 0.130806 | 0.999303 | 0.999072 |
| chrX:108570703:A>C | A>C | High Impact | 2.072266 | 0.999897 | 0.999834 | 0.147736 | 0.12648 | 0.99927 | 0.999015 |
| chrX:108570719:A>C | A>C | High Impact | 1.983398 | 0.999882 | 0.999833 | 0.141876 | 0.123032 | 0.999218 | 0.998988 |
| chrX:108570672:T>G | T>G | High Impact | 1.988281 | 0.999882 | 0.99621 | 0.126801 | 0.108604 | 0.999015 | 0.998778 |
| chrX:108570675:T>G | T>G | High Impact | 1.97168 | 0.999879 | 0.994799 | 0.139923 | 0.119614 | 0.999199 | 0.998935 |
| chrX:108570727:A>G | A>G | High Impact | 1.813477 | 0.999844 | 0.999785 | 0.130432 | 0.112885 | 0.999081 | 0.998876 |

**Table S7.** The list of high spliceogenic substitutions from ISM analysis in intron 1 hotspot.

| **Variant ID** | **Change** | **Impact Classification** | **Overall Max Raw** | **Splice Junction Max Quantile** | **Splice Junction Mean Quantile** | **Splice Site Max Raw** | **Splice Site Mean Raw** | **Splice Site Max Quantile** | **Splice Site Mean Quantile** |
| --- | --- | --- | --- | --- | --- | --- | --- | --- | --- |
| **chrX:108536057:T>A** | T>A | High Impact | 3.939453 | 0.999993 | 0.999874 | 0.78717 | 0.73053 | 0.999976 | 0.999969 |
| **chrX:108535989:A>T** | A>T | High Impact | 3.376953 | 0.999988 | 0.999826 | 0.64917 | 0.588623 | 0.999957 | 0.999947 |
| **chrX:108535992:A>T** | A>T | High Impact | 3.3125 | 0.999987 | 0.999818 | 0.654419 | 0.598022 | 0.999959 | 0.999949 |
| **chrX:108536044:T>C** | T>C | High Impact | 3.083984 | 0.999984 | 0.999796 | 0.562256 | 0.501709 | 0.999938 | 0.999926 |
| **chrX:108536052:T>A** | T>A | High Impact | 2.806641 | 0.999978 | 0.999766 | 0.374451 | 0.333252 | 0.999878 | 0.999859 |
| **chrX:108535980:G>C** | G>C | High Impact | 2.695312 | 0.999975 | 0.999763 | 0.485046 | 0.435242 | 0.999919 | 0.999906 |
| **chrX:108535993:A>T** | A>T | High Impact | 2.654297 | 0.999974 | 0.999757 | 0.440308 | 0.393433 | 0.999904 | 0.999889 |
| **chrX:108536009:T>G** | T>G | High Impact | 2.611328 | 0.999972 | 0.999748 | 0.459473 | 0.40918 | 0.999913 | 0.999898 |
| **chrX:108535978:T>A** | T>A | High Impact | 2.587891 | 0.999972 | 0.999762 | 0.355713 | 0.318359 | 0.99987 | 0.999849 |
| **chrX:108536047:G>A** | G>A | High Impact | 2.427734 | 0.999965 | 0.999701 | 0.287354 | 0.255859 | 0.999829 | 0.999798 |
| **chrX:108536038:T>C** | T>C | High Impact | 2.353516 | 0.999963 | 0.999691 | 0.363892 | 0.320251 | 0.999874 | 0.999851 |
| **chrX:108536006:T>G** | T>G | High Impact | 2.320312 | 0.99996 | 0.999685 | 0.358154 | 0.319336 | 0.99987 | 0.999849 |
| **chrX:108535976:C>A** | C>A | High Impact | 2.263672 | 0.999957 | 0.9997 | 0.4021 | 0.360779 | 0.99989 | 0.999873 |
| **chrX:108536020:A>G** | A>G | High Impact | 2.134766 | 0.99995 | 0.999598 | 0.285217 | 0.253967 | 0.999829 | 0.999798 |
| **chrX:108536039:T>C** | T>C | High Impact | 2.064453 | 0.999944 | 0.999615 | 0.243835 | 0.217255 | 0.999783 | 0.999749 |
| **chrX:108536004:T>G** | T>G | High Impact | 1.996094 | 0.999938 | 0.999591 | 0.257568 | 0.228973 | 0.999797 | 0.999766 |
| **chrX:108536048:T>A** | T>A | High Impact | 1.960938 | 0.999936 | 0.999579 | 0.260986 | 0.232178 | 0.999804 | 0.999774 |
| **chrX:108536045:G>C** | G>C | High Impact | 1.96582 | 0.999936 | 0.999581 | 0.305054 | 0.269226 | 0.99984 | 0.999812 |
| **chrX:108535980:G>A** | G>A | High Impact | 1.864258 | 0.999927 | 0.99957 | 0.246765 | 0.221375 | 0.999783 | 0.999754 |
| **chrX:108536024:T>G** | T>G | High Impact | 1.847656 | 0.999924 | 0.99952 | 0.272583 | 0.243347 | 0.999817 | 0.999788 |
| **chrX:108535989:A>C** | A>C | High Impact | 1.808594 | 0.999919 | 0.999517 | 0.199951 | 0.178955 | 0.999716 | 0.999671 |
| **chrX:108535980:G>T** | G>T | High Impact | 1.789062 | 0.999919 | 0.999535 | 0.23407 | 0.210632 | 0.999768 | 0.999737 |
| **chrX:108536004:T>C** | T>C | High Impact | 1.685547 | 0.999904 | 0.999451 | 0.190186 | 0.17038 | 0.999696 | 0.999648 |
| **chrX:108536023:T>G** | T>G | High Impact | 1.632812 | 0.999897 | 0.999435 | 0.154053 | 0.13974 | 0.999587 | 0.999532 |
| **chrX:108536072:G>T** | G>T | High Impact | 1.570312 | 0.999886 | 0.999432 | 0.202515 | 0.179626 | 0.999725 | 0.999676 |
| **chrX:108536017:T>G** | T>G | High Impact | 1.554688 | 0.999882 | 0.999389 | 0.188965 | 0.16864 | 0.999696 | 0.999642 |
| **chrX:108536046:G>C** | G>C | High Impact | 1.388672 | 0.999846 | 0.999288 | 0.114929 | 0.103943 | 0.99938 | 0.999284 |
| **chrX:108536025:A>C** | A>C | High Impact | 1.34375 | 0.999835 | 0.999253 | 0.130127 | 0.116699 | 0.999477 | 0.999396 |
| **chrX:108536048:T>C** | T>C | High Impact | 1.267578 | 0.999811 | 0.999151 | 0.108643 | 0.0979 | 0.999314 | 0.999223 |
| **chrX:108536061:A>T** | A>T | High Impact | 1.271484 | 0.999811 | 0.99928 | 0.133057 | 0.118835 | 0.999494 | 0.999404 |
| **chrX:108536014:A>G** | A>G | High Impact | 1.242188 | 0.999811 | 0.999143 | 0.093872 | 0.085327 | 0.99916 | 0.999065 |
| **chrX:108536023:T>C** | T>C | High Impact | 1.235352 | 0.999804 | 0.999116 | 0.082764 | 0.075775 | 0.999005 | 0.998893 |
| **chrX:108536072:G>C** | G>C | High Impact | 1.192383 | 0.999783 | 0.999105 | 0.117554 | 0.105164 | 0.999401 | 0.999294 |
| **chrX:108536037:T>G** | T>G | High Impact | 1.105469 | 0.99976 | 0.998979 | 0.112183 | 0.10144 | 0.999359 | 0.999273 |
| **chrX:108536073:G>T** | G>T | High Impact | 1.013672 | 0.999706 | 0.99892 | 0.086914 | 0.077576 | 0.99907 | 0.998925 |
| **chrX:108535981:A>T** | A>T | High Impact | 0.995117 | 0.999696 | 0.998889 | 0.085083 | 0.077271 | 0.999038 | 0.99893 |
| **chrX:108536072:G>A** | G>A | High Impact | 0.935059 | 0.999652 | 0.998784 | 0.082886 | 0.075134 | 0.999005 | 0.998893 |

**Table S8.** The list of high spliceogenic substitutions from ISM analysis in intron 4 hotspot.

| **Variant ID** | **Change** | **Impact Classification** | **Overall Max Raw** | **Splice Junction Max Quantile** | **Splice Junction Mean Quantile** | **Splice Site Max Raw** | **Splice Site Mean Raw** | **Splice Site Max Quantile** | **Splice Site Mean Quantile** |
| --- | --- | --- | --- | --- | --- | --- | --- | --- | --- |
| **chrX:108568096:T>G** | T>G | High Impact | 6.980469 | 1 | 0.999999 | 0.912605 | 0.875519 | 0.999996 | 0.99999 |
| **chrX:108568071:G>C** | G>C | High Impact | 5.1875 | 0.999998 | 0.999994 | 0.873451 | 0.814926 | 0.99999 | 0.999981 |
| **chrX:108568056:A>C** | A>C | High Impact | 5.015625 | 0.999997 | 0.999855 | 0.829407 | 0.759247 | 0.999984 | 0.999974 |
| **chrX:108568068:C>G** | C>G | High Impact | 4.84375 | 0.999997 | 0.998409 | 0.613266 | 0.541977 | 0.999951 | 0.999936 |
| **chrX:108568070:A>C** | A>C | High Impact | 4.863281 | 0.999997 | 0.999793 | 0.802292 | 0.732403 | 0.999978 | 0.99997 |
| **chrX:108568069:T>G** | T>G | High Impact | 4.589844 | 0.999996 | 0.999748 | 0.755096 | 0.675179 | 0.999971 | 0.99996 |
| **chrX:108568069:T>C** | T>C | High Impact | 3.824219 | 0.999992 | 0.999988 | 0.548065 | 0.471214 | 0.999936 | 0.999917 |
| **chrX:108568069:T>A** | T>A | High Impact | 3.662109 | 0.999991 | 0.999986 | 0.512909 | 0.440941 | 0.999924 | 0.999905 |
| **chrX:108568070:A>G** | A>G | High Impact | 3.609375 | 0.99999 | 0.999985 | 0.484734 | 0.42128 | 0.999919 | 0.9999 |
| **chrX:108568056:A>G** | A>G | High Impact | 2.833984 | 0.999978 | 0.999972 | 0.315735 | 0.27179 | 0.999846 | 0.999811 |
| **chrX:108568048:A>G** | A>G | High Impact | 2.755859 | 0.999977 | 0.99997 | 0.285461 | 0.248119 | 0.999829 | 0.99979 |
| **chrX:108568090:G>A** | G>A | High Impact | 2.648438 | 0.999974 | 0.999968 | 0.241463 | 0.205917 | 0.999783 | 0.999723 |
| **chrX:108568043:A>C** | A>C | High Impact | 2.628906 | 0.999972 | 0.999968 | 0.221199 | 0.193512 | 0.999752 | 0.999702 |
| **chrX:108568026:A>T** | A>T | High Impact | 2.582031 | 0.999972 | 0.999966 | 0.23745 | 0.20945 | 0.999776 | 0.999736 |
| **chrX:108568070:A>T** | A>T | High Impact | 2.449219 | 0.999966 | 0.999962 | 0.267937 | 0.234756 | 0.999811 | 0.999773 |
| **chrX:108568089:T>C** | T>C | High Impact | 2.40625 | 0.999964 | 0.999957 | 0.205826 | 0.181431 | 0.999725 | 0.999676 |
| **chrX:108568077:A>C** | A>C | High Impact | 2.285156 | 0.999959 | 0.999952 | 0.280884 | 0.246735 | 0.999823 | 0.999792 |
| **chrX:108568023:A>T** | A>T | High Impact | 2.242188 | 0.999956 | 0.999802 | 0.173798 | 0.151993 | 0.999652 | 0.999573 |
| **chrX:108568048:A>C** | A>C | High Impact | 2.228516 | 0.999956 | 0.999952 | 0.176086 | 0.151257 | 0.999663 | 0.99957 |
| **chrX:108568055:T>G** | T>G | High Impact | 2.169922 | 0.999951 | 0.999946 | 0.152542 | 0.131207 | 0.999587 | 0.999484 |
| **chrX:108568090:G>C** | G>C | High Impact | 2.035156 | 0.999942 | 0.999936 | 0.115974 | 0.101887 | 0.99938 | 0.999256 |
| **chrX:108568065:A>C** | A>C | High Impact | 2.003906 | 0.99994 | 0.999932 | 0.110001 | 0.09808 | 0.999337 | 0.999234 |
| **chrX:108568043:A>G** | A>G | High Impact | 1.910156 | 0.999931 | 0.999929 | 0.088875 | 0.079254 | 0.999101 | 0.998961 |
| **chrX:108568063:A>G** | A>G | High Impact | 1.892578 | 0.999929 | 0.999925 | 0.099426 | 0.08585 | 0.999215 | 0.999057 |
| **chrX:108568055:T>C** | T>C | High Impact | 1.858398 | 0.999927 | 0.999921 | 0.09639 | 0.083286 | 0.999188 | 0.999005 |
| **chrX:108568078:G>C** | G>C | High Impact | 1.799805 | 0.999919 | 0.999919 | 0.126686 | 0.113991 | 0.999459 | 0.999375 |
| **chrX:108568104:G>T** | G>T | High Impact | 1.791992 | 0.999919 | 0.999916 | 0.107742 | 0.093708 | 0.999314 | 0.99916 |

**Table S9.** The list of high impact spliceogenic substitutions from ISM analysis in intron 30 hotspot.

| **Variant ID** | **Change** | **Impact Classification** | **Overall Max Raw** | **Splice Junction Max Quantile** | **Splice Junction Mean Quantile** | **Splice Site Max Raw** | **Splice Site Mean Raw** | **Splice Site Max Quantile** | **Splice Site Mean Quantile** |
| --- | --- | --- | --- | --- | --- | --- | --- | --- | --- |
| **chrX:108615623:A>T** | A>T | High Impact | 6.75 | 1 | 0.998894 | 0.816377 | 0.796843 | 0.999982 | 0.999979 |
| **chrX:108615623:A>C** | A>C | High Impact | 5.832031 | 0.999999 | 0.999998 | 0.636689 | 0.611296 | 0.999956 | 0.999952 |
| **chrX:108615624:G>T** | G>T | High Impact | 5.734375 | 0.999999 | 0.999998 | 0.578102 | 0.550756 | 0.999942 | 0.999938 |
| **chrX:108615627:A>C** | A>C | High Impact | 5.367188 | 0.999998 | 0.999529 | 0.835083 | 0.811573 | 0.999985 | 0.999981 |
| **chrX:108615610:G>T** | G>T | High Impact | 5.257812 | 0.999998 | 0.999384 | 0.749393 | 0.721529 | 0.99997 | 0.999967 |
| **chrX:108615622:T>C** | T>C | High Impact | 5.007812 | 0.999997 | 0.999418 | 0.731979 | 0.706429 | 0.999969 | 0.999966 |
| **chrX:108615624:G>C** | G>C | High Impact | 4.992188 | 0.999997 | 0.999996 | 0.457008 | 0.435522 | 0.999913 | 0.999909 |
| **chrX:108615628:G>C** | G>C | High Impact | 4.722656 | 0.999997 | 0.999403 | 0.545807 | 0.512516 | 0.999934 | 0.999929 |
| **chrX:108615627:A>T** | A>T | High Impact | 4.613281 | 0.999996 | 0.999473 | 0.590469 | 0.557106 | 0.999944 | 0.999939 |
| **chrX:108615647:T>G** | T>G | High Impact | 4.464844 | 0.999996 | 0.999432 | 0.636292 | 0.608734 | 0.999956 | 0.999952 |
| **chrX:108615625:A>G** | A>G | High Impact | 4.398438 | 0.999995 | 0.999183 | 0.376244 | 0.352747 | 0.999882 | 0.999871 |
| **chrX:108615657:T>C** | T>C | High Impact | 4.160156 | 0.999994 | 0.999246 | 0.467838 | 0.441393 | 0.999916 | 0.99991 |
| **chrX:108615611:G>T** | G>T | High Impact | 4.1875 | 0.999994 | 0.999401 | 0.546951 | 0.520332 | 0.999934 | 0.999931 |
| **chrX:108615659:A>G** | A>G | High Impact | 3.861328 | 0.999992 | 0.999323 | 0.515076 | 0.48848 | 0.999927 | 0.999923 |
| **chrX:108615615:G>T** | G>T | High Impact | 3.769531 | 0.999992 | 0.999268 | 0.444183 | 0.424454 | 0.999907 | 0.999904 |
| **chrX:108615610:G>C** | G>C | High Impact | 3.623047 | 0.99999 | 0.99891 | 0.359871 | 0.343174 | 0.999874 | 0.999867 |
| **chrX:108615616:C>T** | C>T | High Impact | 3.449219 | 0.999989 | 0.99888 | 0.346451 | 0.333675 | 0.999865 | 0.999863 |
| **chrX:108615637:A>C** | A>C | High Impact | 3.462891 | 0.999989 | 0.999085 | 0.34063 | 0.325901 | 0.99986 | 0.999856 |
| **chrX:108615598:C>G** | C>G | High Impact | 3.417969 | 0.999988 | 0.999987 | 0.139647 | 0.137693 | 0.999528 | 0.999528 |
| **chrX:108615641:T>A** | T>A | High Impact | 3.339844 | 0.999987 | 0.998891 | 0.244148 | 0.234787 | 0.999783 | 0.999779 |
| **chrX:108615627:A>G** | A>G | High Impact | 3.304688 | 0.999987 | 0.999049 | 0.346329 | 0.325661 | 0.999865 | 0.999858 |
| **chrX:108615683:T>G** | T>G | High Impact | 2.876953 | 0.99998 | 0.999075 | 0.286804 | 0.272934 | 0.999829 | 0.99982 |
| **chrX:108615628:G>T** | G>T | High Impact | 2.814453 | 0.999978 | 0.998072 | 0.159088 | 0.153141 | 0.999601 | 0.999594 |
| **chrX:108615647:T>C** | T>C | High Impact | 2.751953 | 0.999977 | 0.998207 | 0.284729 | 0.272797 | 0.999823 | 0.999817 |
| **chrX:108615657:T>G** | T>G | High Impact | 2.722656 | 0.999976 | 0.998482 | 0.142643 | 0.138659 | 0.999543 | 0.999527 |
| **chrX:108615619:C>T** | C>T | High Impact | 2.685547 | 0.999975 | 0.998268 | 0.255157 | 0.246628 | 0.999797 | 0.999794 |
| **chrX:108615659:A>C** | A>C | High Impact | 2.623047 | 0.999972 | 0.998804 | 0.263123 | 0.250198 | 0.999804 | 0.999794 |
| **chrX:108615668:T>C** | T>C | High Impact | 2.511719 | 0.999969 | 0.998513 | 0.139656 | 0.135178 | 0.999528 | 0.999511 |
| **chrX:108615615:G>C** | G>C | High Impact | 2.515625 | 0.999969 | 0.997921 | 0.174652 | 0.170547 | 0.999663 | 0.999658 |
| **chrX:108615668:T>G** | T>G | High Impact | 2.482422 | 0.999967 | 0.998463 | 0.128914 | 0.126877 | 0.999477 | 0.999468 |
| **chrX:108615663:C>G** | C>G | High Impact | 2.291016 | 0.999959 | 0.997637 | 0.116592 | 0.113983 | 0.99938 | 0.99938 |
| **chrX:108615674:T>C** | T>C | High Impact | 2.236328 | 0.999956 | 0.997867 | 0.093712 | 0.092365 | 0.999188 | 0.999159 |
| **chrX:108615711:T>G** | T>G | High Impact | 2.199219 | 0.999953 | 0.997262 | 0.117386 | 0.115425 | 0.999401 | 0.999401 |
| **chrX:108615702:A>G** | A>G | High Impact | 2.154297 | 0.999951 | 0.997345 | 0.107544 | 0.105927 | 0.999314 | 0.999314 |
| **chrX:108615635:A>G** | A>G | High Impact | 2.119141 | 0.999948 | 0.997569 | 0.126282 | 0.123611 | 0.999459 | 0.99945 |
| **chrX:108615648:A>G** | A>G | High Impact | 2.072266 | 0.999944 | 0.997649 | 0.091316 | 0.090462 | 0.99916 | 0.999145 |
| **chrX:108615611:G>C** | G>C | High Impact | 2.001953 | 0.99994 | 0.997006 | 0.106033 | 0.104317 | 0.999314 | 0.999302 |
| **chrX:108615683:T>C** | T>C | High Impact | 1.994141 | 0.999938 | 0.997734 | 0.110046 | 0.106918 | 0.999337 | 0.999326 |
| **chrX:108615711:T>C** | T>C | High Impact | 1.958984 | 0.999936 | 0.996883 | 0.089066 | 0.088814 | 0.999131 | 0.999116 |
| **chrX:108615710:T>C** | T>C | High Impact | 1.808594 | 0.999922 | 0.997702 | 0.11908 | 0.116837 | 0.999401 | 0.999401 |
| **chrX:108615635:A>C** | A>C | High Impact | 1.760742 | 0.999916 | 0.996739 | 0.08136 | 0.080643 | 0.999005 | 0.998988 |

**Table S10.** The list of high impact spliceogenic substitutions from ISM analysis in intron 44 hotspot.

| **Variant ID** | **Change** | **Impact Classification** | **Overall Max Raw** | **Splice Junction Max Quantile** | **Splice Junction Mean Quantile** | **Splice Site Max Raw** | **Splice Site Mean Raw** | **Splice Site Max Quantile** | **Splice Site Mean Quantile** |
| --- | --- | --- | --- | --- | --- | --- | --- | --- | --- |
| chrX:108680214:A>T | A>T | High Impact | 5.132812 | 0.999998 | 0.998699 | 0.659454 | 0.656448 | 0.999962 | 0.99996 |
| chrX:108680215:G>T | G>T | High Impact | 3.6875 | 0.999991 | 0.998304 | 0.338379 | 0.323753 | 0.99987 | 0.999858 |
| chrX:108680214:A>C | A>C | High Impact | 3.667969 | 0.999991 | 0.996947 | 0.352814 | 0.337112 | 0.999878 | 0.999864 |
| chrX:108680180:A>G | A>G | High Impact | 3.203125 | 0.999987 | 0.999045 | 0.41861 | 0.40303 | 0.999904 | 0.999895 |
| chrX:108680234:T>C | T>C | High Impact | 3.128906 | 0.999984 | 0.998621 | 0.328613 | 0.313011 | 0.99986 | 0.999848 |
| chrX:108680250:A>C | A>C | High Impact | 2.494141 | 0.999975 | 0.999972 | 0.175354 | 0.160583 | 0.999674 | 0.999616 |
| chrX:108680215:G>C | G>C | High Impact | 2.488281 | 0.999971 | 0.996023 | 0.094238 | 0.084007 | 0.999215 | 0.999018 |
| chrX:108680239:T>G | T>G | High Impact | 2.449219 | 0.999969 | 0.998233 | 0.151916 | 0.137894 | 0.999601 | 0.999521 |
| chrX:108680248:A>G | A>G | High Impact | 2.253906 | 0.999963 | 0.996383 | 0.169678 | 0.155548 | 0.999663 | 0.999603 |
| chrX:108680262:T>C | T>C | High Impact | 2.103516 | 0.999962 | 0.999955 | 0.101273 | 0.08979 | 0.999291 | 0.999113 |
| chrX:108680238:T>G | T>G | High Impact | 2.109375 | 0.999954 | 0.980056 | 0.08197 | 0.072296 | 0.999038 | 0.998821 |

**Table S11.** The list of high impact spliceogenic substitutions from ISM analysis in intron 49 hotspot.

| **Variant ID** | **Change** | **Impact Classification** | **Overall Max Raw** | **Splice Junction Max Quantile** | **Splice Junction Mean Quantile** | **Splice Site Max Raw** | **Splice Site Mean Raw** | **Splice Site Max Quantile** | **Splice Site Mean Quantile** |
| --- | --- | --- | --- | --- | --- | --- | --- | --- | --- |
| **chrX:108688887:C>G** | C>G | High Impact | 4.640625 | 0.999996 | 0.999762 | 0.557987 | 0.542263 | 0.999938 | 0.999936 |
| **chrX:108688919:T>G** | T>G | High Impact | 4.039062 | 0.999993 | 0.999702 | 0.475151 | 0.470985 | 0.999922 | 0.999919 |
| **chrX:108688968:G>T** | G>T | High Impact | 3.792969 | 0.999992 | 0.999991 | 0.354614 | 0.349855 | 0.999878 | 0.999872 |
| **chrX:108688961:G>A** | G>A | High Impact | 3.466797 | 0.999989 | 0.999272 | 0.17783 | 0.173065 | 0.999685 | 0.999663 |
| **chrX:108688951:T>C** | T>C | High Impact | 3.083984 | 0.999984 | 0.999545 | 0.350891 | 0.349487 | 0.999874 | 0.99987 |
| **chrX:108688910:T>G** | T>G | High Impact | 3.058594 | 0.999984 | 0.999445 | 0.165161 | 0.159904 | 0.999652 | 0.99962 |
| **chrX:108688903:G>C** | G>C | High Impact | 2.951172 | 0.999983 | 0.998979 | 0.14175 | 0.136499 | 0.999558 | 0.999518 |
| **chrX:108688915:T>G** | T>G | High Impact | 2.935547 | 0.999982 | 0.999121 | 0.157204 | 0.151978 | 0.999627 | 0.999593 |
| **chrX:108688905:T>A** | T>A | High Impact | 2.849609 | 0.999981 | 0.999104 | 0.109177 | 0.104141 | 0.999359 | 0.999287 |
| **chrX:108688940:C>G** | C>G | High Impact | 2.849609 | 0.99998 | 0.999452 | 0.125122 | 0.119377 | 0.999477 | 0.999418 |
| **chrX:108688917:C>G** | C>G | High Impact | 2.732422 | 0.999978 | 0.999116 | 0.154068 | 0.148859 | 0.999614 | 0.999579 |
| **chrX:108688875:A>T** | A>T | High Impact | 2.494141 | 0.999977 | 0.999359 | 0.094696 | 0.08915 | 0.999215 | 0.999126 |
| **chrX:108688930:G>C** | G>C | High Impact | 2.636719 | 0.999976 | 0.998985 | 0.110329 | 0.105335 | 0.99938 | 0.999311 |
| **chrX:108688872:G>T** | G>T | High Impact | 2.453125 | 0.999974 | 0.999337 | 0.084408 | 0.079844 | 0.99907 | 0.998965 |
| **chrX:108688918:C>G** | C>G | High Impact | 2.527344 | 0.999972 | 0.999289 | 0.109962 | 0.105751 | 0.99938 | 0.999311 |
| **chrX:108688941:A>G** | A>G | High Impact | 2.392578 | 0.99997 | 0.999078 | 0.083717 | 0.079977 | 0.99907 | 0.998984 |

**Table S12.** Significant motifs identified by MSEA of spliceogenic substitutions in intron 6 hotspot

| Description | setSize | enrichmentScore | NES | pvalue | p.adjust | qvalue | rank | core_enrichment |
| --- | --- | --- | --- | --- | --- | --- | --- | --- |
| CG-core | 79 | 0.41174651 | 3.34562511 | 1.19E-09 | 7.87E-08 | 6.15E-08 | 113 | TTCGAGG/AAGCGTA/AGTCGTA/GAACGGT/TTCGGAT/AGACGGG/CAACGAC/CACGCAA/AACGGTA/TTCGAAC/CCGCGGC/CAACGTG/CTCGCAA/CACGAAG/TCTCGGA/CTACGAT/AGTCGTT/TCCGTTC/TTCGCTC/ATTCGAC/GGCGACT/ACCGACT/TTCGTTT/GACGCAC/TACGTTT/CTCGCGG/CACGTCC/TATCGGA/CGTCGCA/TTCGATG/AGGCGGA/CCACGTG/GAGCGAA/GCCGCTT/TCTCGGC/TTCGTTC/ACACGGT/CACGACA/ATGCGTA/CGTAGCA/ACCGTCT/AACGAGG/CTCCGGA |
| TCG | 26 | 0.50902005 | 2.77464752 | 5.98E-06 | 0.00019736 | 0.00015424 | 86 | TTCGAGG/AGTCGTA/TTCGGAT/TTCGAAC/CTCGCAA/TCTCGGA/AGTCGTT/TTCGCTC/ATTCGAC/TTCGTTT/CTCGCGG/TATCGGA/CGTCGCA/TTCGATG/TCTCGGC/TTCGTTC |
| CGC | 10 | 0.66472816 | 2.43558782 | 0.00017336 | 0.00381386 | 0.00298053 | 83 | CACGCAA/CCGCGGC/CTCGCAA/TTCGCTC/GACGCAC/CTCGCGG/CGTCGCA/GCCGCTT |
| ACG | 25 | 0.42000849 | 2.26116298 | 0.000715 | 0.01179758 | 0.0092198 | 111 | GAACGGT/AGACGGG/CAACGAC/CACGCAA/AACGGTA/CAACGTG/CACGAAG/CTACGAT/GACGCAC/TACGTTT/CACGTCC/CCACGTG/ACACGGT/CACGACA/AACGAGG |
| CGG | 19 | 0.45234955 | 2.16853256 | 0.00180512 | 0.0238276 | 0.01862125 | 113 | GAACGGT/TTCGGAT/AGACGGG/AACGGTA/CCGCGGC/TCTCGGA/CTCGCGG/TATCGGA/AGGCGGA/TCTCGGC/ACACGGT/CTCCGGA |
| CGA | 21 | 0.41387152 | 2.07390354 | 0.00274879 | 0.03023668 | 0.02362994 | 111 | TTCGAGG/CAACGAC/TTCGAAC/CACGAAG/CTACGAT/ATTCGAC/GGCGACT/ACCGACT/TTCGATG/GAGCGAA/CACGACA/AACGAGG |
| CGT | 27 | 0.37341952 | 2.08291142 | 0.00419906 | 0.03959113 | 0.03094044 | 269 | AAGCGTA/AGTCGTA/CAACGTG/AGTCGTT/TCCGTTC/TTCGTTT/TACGTTT/CACGTCC/CGTCGCA/CCACGTG/TTCGTTC/ATGCGTA/CGTAGCA/ACCGTCT/TCCGTGT/AACCGTA/TTAGCGT/TTAACGT/TCGTGAC/CGTTGCA/GACGTAC/AACGTTC/TGACGTT/TGACGTC |

**Table S13.** List of all primers and their sequences used in this study.

| Primers | Aim | Strand | Sequence (5'-3') |
| --- | --- | --- | --- |
| c.385-756C>T | Mutagenesis | Forward | AGGCCACTTCtTTTCTCTCGG |
|  |  | Reverse | AGACATCAAGTTTAAAAGACAGAAAAAG |
| c.385-679T>C | Mutagenesis | Forward | CACACAACAGcAGTAATTTTATTCTTTC |
|  |  | Reverse | TCTTTGTGAGTCCAATAC |
| c.385-660A>G | Mutagenesis | Forward | TATTCTTTCAgACCTTCTGATGAAG |
|  |  | Reverse | AAATTACTACTGTTGTGTGTC |
| c.385-719G>A | Mutagenesis | Forward | GTGTAACAAGaTGGAGAGAAG |
|  |  | Reverse | ATGGAAAAAGTAGGTCCC |
| c.385-716G>A | Mutagenesis | Forward | TAACAAGGTGaAGAGAAGGGTATTG |
|  |  | Reverse | CACATGGAAAAAGTAGGTC |
| c.385-714 G>A | Mutagenesis | Forward | ACAAGGTGGAaAGAAGGGTAT |
|  |  | Reverse | TACACATGGAAAAAGTAGGTC |
| c.385-756C>G | Mutagenesis | Forward | AGGCCACTTCgTTTCTCTCGG |
|  |  | Reverse | AGACATCAAGTTTAAAAGACAGAAAAAG |
| c.385-752C>G | Mutagenesis | Forward | CACTTCCTTTgTCTCGGGACC |
|  |  | Reverse | GCCTAGACATCAAGTTTAAAAG |
| c.385-708G>C | Mutagenesis | Forward | TGGAGAGAAGcGTATTGGACTC |
|  |  | Reverse | CCTTGTTACACATGGAAAAAG |
| c.385-708G>A | Mutagenesis | Forward | TGGAGAGAAGaGTATTGGACTC |
|  |  | Reverse | CCTTGTTACACATGGAAAAAG |
| Pspl3_COL4A5_Int4_F | SNAP Assembly | Forward | TTATGGGGTACGGGATCACCAGAATTATGAAGACAATGCTCCCTCA |
| COL4A5_Int6_R1 |  | Reverse | ATAGCCAAGACCTGAAGACA |
| Hom_COL4A5_Int6_F1 | SNAP Assembly | Forward | TGTCTTCAGGTCTTGGCTATGATTAACTTGTGTTTCAGGCT |
| Hom_COL4A5_int6_R2 |  | Reverse | AGATGGTAAGAAAGAAGGCCA |
| Hom_COL4A5_Int6_F2 | SNAP Assembly | Forward | TGGCCTTCTTTCTTACCATCTGCAATCTGGCATGTTTCAAAATG |
| Pspl3_COL4A5_Int7_R |  | Reverse | GGCCGCTCGAGCTCCAGAATTTTTTGCCCAGGAGAAAGCTA |
| Pspl3_V1_F | RT-PCR | Forward | TCTGAGTCACCTGGACAACC |
| Pspl3_V2_R |  | Reverse | ATCTCAGTGGTATTTGTGAGC |
| Pspl3_pseudoexon_F | RT-PCR | Forward | GGACTCACAAAGACACACAACA |
| *COL4A5*_RT_PCR | RT-PCR | Forward | CACCAGGACCAAAAGGAATCA |
|  |  | Reverse | ACCAGGAAAACCGGGACTG |
| *COL4A5*_qPCR | QPCR | Forward | CAGACGATCCAGATTCCCCATTGT |
|  |  | Reverse | GAACCAGGGGAGGCTAGGGCTTGA |
| *HPRT1* | QPCR | Forward | TCTTTGCTGACCTGCTGGATT |
|  |  | Reverse | GTTGAGAGATCATCTCCACCAATTACT |
